# Mental health outcomes following COVID-19 infection: Evidence from 11 UK longitudinal population studies

**DOI:** 10.1101/2022.05.11.22274964

**Authors:** Ellen J. Thompson, Jean Stafford, Bettina Moltrecht, Charlotte F. Huggins, Alex S. F. Kwong, Richard J. Shaw, Paola Zaninotto, Kishan Patel, Richard J. Silverwood, Eoin McElroy, Matthias Pierce, Michael J. Green, Ruth C. E. Bowyer, Jane Maddock, Kate Tilling, S. Vittal Katikireddi, George B. Ploubidis, David J. Porteous, Nic Timpson, Nish Chaturvedi, Claire J. Steves, Praveetha Patalay

**Affiliations:** Department of Twin Research and Genetic Epidemiology, School of Life Course & Population Sciences, King’s College London; MRC Unit for Lifelong Health and Ageing, University College London (UCL); Centre for Longitudinal Studies, UCL; Centre for Genomic and Experimental Medicine, University of Edinburgh; Division of Psychiatry, University of Edinburgh; MRC/CSO Social & Public Health Sciences Unit, Institute of Health & Wellbeing, University of Glasgow; Department of Epidemiology and Public Health, UCL; School of Psychology, Ulster University; Division of Psychology & Mental Health, The University of Manchester; Population Health Sciences, Bristol Medical School, University of Bristol

**Author notes:** Joint first authors (Ellen J. Thompson, Jean Stafford, Bettina Moltrecht).

**Keywords:** SARS CoV-2, COVID-19, psychological distress, depression, anxiety

## Abstract

**Background:** Evidence on associations between COVID-19 illness and mental health is mixed. We examined longitudinal associations between COVID-19 and mental health while considering: 1) pre-pandemic mental health, 2) time since infection; 3) subgroup differences; and 4) confirmation of infection via self-reported test, and serology data.

**Methods:** Using data from 11 UK longitudinal studies, involving 54,442 participants, with 2 to 8 repeated measures of mental health and COVID-19 between April 2020 and April 2021, we standardised continuous mental health scales within each study across time. We investigated associations between COVID-19 (self-report, test-confirmed, serology-confirmed) and mental health using multilevel generalised estimating equations. We examined whether associations varied by age, sex, ethnicity, education and pre-pandemic mental health. Effect-sizes were pooled in random-effects meta-analyses.

**Outcomes:** Pooled estimates of the standardized difference in outcome between those with and without self-reported COVID-19 suggested associations with subsequent psychological distress (0.10 [95%CI: 0.06; 0.13], I^2^=42.8%), depression (0.08 [0.05; 0.10], I^2^=20.8%), anxiety (0.08 [0.05; 0.10], I^2^=0%), and lower life satisfaction (−0.06 [-0.08; -0.04], I^2^=29.2%). Associations did not vary by time since infection until 3+ months and were present in all age groups, with some evidence of stronger effects in those aged 50+. Self-reported COVID-19, whether suspected or test-confirmed and irrespective of serology status, was associated with poorer mental health.

**Interpretation:** Self-reporting COVID-19 was longitudinally associated with deterioration in mental health and life satisfaction. Our findings have important implications for mental health service provision, given the substantial prevalence of COVID-19 in the UK and worldwide.

**Funding:** MRC and NIHR

## Introduction

Infection with severe acute respiratory syndrome coronavirus 2 (SARS-CoV-2) can cause either asymptomatic or symptomatic coronavirus disease 2019 (COVID-19). Mental ill-health is increasingly recognised as a potential consequence of COVID-19, following initial evidence from case reports^1^ and studies of other severe coronavirus infections.^2^ However, longitudinal evidence in this area is limited and few studies have sought to disentangle the effects of COVID-19 illness from the wider mental health impacts of the pandemic. As such, the mental health consequences of COVID-19 in the general population remain poorly understood.

Recent systematic reviews have yielded mixed results as to whether COVID-19 illness is associated with psychological distress,^3,4^ which may reflect a lack of high-quality longitudinal evidence in this area. Previous studies have been limited by small and/or unrepresentative samples, cross-sectional designs and absence of control groups.^5,6^ Although several studies using routine data reported elevated rates of psychiatric disorders following COVID-19 illness,^7–12^ others have not found clear evidence of associations.^13,14^ Further, most studies using routine data lacked detailed information on pre-pandemic health and socio-demographic factors, and mainly focused on more severe COVID-19 and recorded mental health disorders.^15,16^ A study using data from six cohorts in Europe found that severe acute COVID-19 illness was associated with adverse mental health outcomes.^17^ In addition, longitudinal studies in the UK have found associations between COVID-19 and psychological distress,^18–20^ although findings have been mixed across different mental health outcomes^21^ and, in the COVID Symptom Study a modest association was found in older participants only.^18^ Further longitudinal research is needed to clarify previous mixed findings, to investigate the magnitude of any association, and to examine whether associations are sustained in the longer-term post-infection.

Using data from 11 UK longitudinal studies we aimed to investigate mental health consequences following COVID-19 illness up to April 2021. First, we examined whether individuals with self-reported COVID-19 experience higher levels of subsequent psychological distress, depression and anxiety, and lower life satisfaction than those without COVID-19. Second, we examined whether associations varied depending on how much time had passed since infection to determine whether effects persist beyond the acute phase of the illness. Third, we explored whether associations varied by age, sex, ethnicity, education and pre-pandemic mental health. Fourth, we examined whether associations between COVID-19 and mental health differed between those with: a) suspected vs test-confirmed COVID-19, and b) self-reported vs serology-detected COVID-19.

## Methods

### Design

The UK National Core Studies – Longitudinal Health and Wellbeing programme combines data from multiple UK population-based longitudinal studies to support more robust inferences that are replicable across data sources. Co-ordinated analysis across different datasets minimises methodological heterogeneity and maximises comparability, while appropriately accounting for study designs and characteristics of individual datasets. Analyses were preregistered (https://osf.io/ntmqw/).

### Participants and sample

Data were drawn from 11 longitudinal UK population studies which conducted surveys before and during the COVID-19 pandemic. For the latter, serology data indicating the presence or absence of a SARS-CoV-2 infection are also available. Details of study designs, timing of the most recent pre-pandemic and COVID-19 surveys, response rates, and analytical sample sizes are shown in Table 1. Ethics statements and data access details are provided in Table S1.

**Table 1.** Details of each study.

| Study Population | Design and Sample Frame | 2020 Age Range | Pre-pandemic Survey | Details of COVID-19 surveys (response rate) | Mental Health Measure | Serology data | Analytic N |
| --- | --- | --- | --- | --- | --- | --- | --- |
| *Age Homogenous Cohorts* |  |  |  |  |  |  |  |
| MCS: Millennium Cohort Study<sup>(1,2)</sup> | Cohort of UK children born between Sept 2000 and Jan 2002 with regular follow-up surveys from birth. | 18-20 | 2018 | Spring 2021 survey response with the issued sample: 33.1% | Kessler-6 (K6) | Taken April –June 2021<br>Valid samples n= 987 | 4652 |
| ALSPAC (G1): Avon Longitudinal Study of Parents and Children-Generation 1 | Cohort of children born in the South-West of England between April 1991 and Dec 1992, with regular follow-up surveys from birth (original young people). | 27-29 | 2017-2018 | Three questionnaires: April (19%), June (17.4%), December (26.4%) | Short Mood and Feelings Questionnaire (SMFQ) | Not available | 2498 |
| NS: Next Steps, formerly known as Longitudinal Study of Young People in England | Sample recruited via secondary schools in England at around age 13 with regular follow-up surveys thereafter. | 29-31 | 2015 | Spring 2021 survey response with the issued sample: 34.3% | General Health Questionnaire 12 (GHQ-12) | Taken April –June 2021<br>Valid samples n= 1037 | 4092 |
| BCS70: British Cohort Study 1970 | Cohort of all children born in Great Britain (i.e., England, Wales & Scotland) in one week in 1970, with regular follow-up surveys from birth. | 50 | 2016 | Spring 2021 survey response with the issued sample: 45.4% | 9-item Malaise inventory | Taken April –June 2021<br>Valid samples n= 2074 | 5545 |
| NCDS: National Child Development Study | Cohort of all children born in Great Britain (i.e., England, Wales & Scotland) in one week in 1958, with regular follow-up surveys from birth. | 62 | 2013 | Spring 2021 survey response with the issued sample: 58.5% | 9-item Malaise inventory | Taken April –June 2021<br>Valid samples n= 2722 | 6696 |
| NSHD: National Survey of Health and Development | Cohort of all children born in Great Britain (i.e. England, Wales & Scotland) in one week in 1946, with regular follow-up surveys from birth | 74 | 2015 | Three surveys: May 2020 (68.2%, Sep-Oct 2020 (61.5%), Feb-Mar 2021 (89.9%) | General Health Questionnaire 12 (GHQ-12) | Taken April –June 2021<br>Valid samples n= 697 | 1721 |
| *Age Heterogeneous Studies* |  |  |  |  |  |  |  |
| USOC: Understanding Society: the UK Household Longitudinal Survey | A nationally representative longitudinal household panel study, based on a clustered-stratified probability sample of UK households, with all adults aged 16+ in chosen households surveyed annually. | 16-96 | 2018-2019 | Seven surveys (full/partial interview): April 2020 (42.0%); May (35.1%); Jun (33.5%); July (32.6%); Sep (30.6%), Nov (28.6%), Jan 2021 (28.5%) | General Health Questionnaire 12 (GHQ-12) | Taken April –June 2021<br>Valid samples n= 6006 | 14154 |
| ELSA: English Longitudinal Study of Aging | A nationally representative population study of individuals aged 50+ living in England, with biennial surveys and periodic refreshing of the sample to maintain representativeness. | 52-90+ | 2018-2019 | Two surveys: Jun-July (75%); Nov-Dec (73%) | Centre for Epidemiological Studies – Depression (CES-D) | Not available | 4752 |
| GS: Generation Scotland: the Scottish Family Health Study(10) | A family-structured, population-based Scottish cohort, with participants aged 18-99 recruited between 2006-2011 | 27-100 | 2006-2011 | Three surveys: April-Jun 2020 (21.3%); Jul-Aug 2020 (15.4%); Feb 2021 (14.3%) | Patient Health Questionnaire 9 (PHQ-9) and Generalised Anxiety Disorder Assessment 7 (GAD-7) | Not available | 3937 |
| ALSPAC(G0): Avon Longitudinal Study of Parents and Children-Generation 0(11) | Parents of the ALSPAC(G1) cohort described above, treated as a separate age-heterogenous study population (original parents). | 45-81 | 2011-2013 | Three questionnaires: April (12.4%), June (12.2%), December (14.3%) | Short Mood and Feelings Questionnaire (SMFQ) | Not available | 3258 |
| TWINSUK: the UK Adult Twin Registry | A cohort of UK volunteer adult twins (55% monozygotic and 43% dizygotic) who were sampled between 18-101 years of age. | 22-96 | 2017-2018 | Three surveys: July 2020 (77.6%), November 2020 (76.1%), March 2021 (76%) | Hospital and Anxiety Depression Scale (HADS) | Taken April 2021<br>Valid samples n= 3137 | 3137 |

Six studies were birth cohorts with all individuals of a similar age: the Millennium Cohort Study (MCS; born 2000-02); the Avon Longitudinal Study of Parents and Children (ALSPAC-G1, born 1990-91); Next Steps (NS, formerly known as the Longitudinal Study of Young People in England; born 1989-90); the 1970 British Cohort Study (BCS70), the National Child Development Study (NCDS; born 1958); and the National Survey of Health and Development (NSHD; born 1946). Five studies were age heterogeneous: Understanding Society/The UK Household Longitudinal Study (USoc/UKHLS); the English Longitudinal Study of Ageing (ELSA); Generation Scotland: The Scottish Family Health Study (GS); the UK Adult Twin Registry (TwinsUK), and the parents of the ALSPAC-G1 birth cohort (ALSPAC-G0).

Analytical samples included participants who had information available on at least one mental health measure, and COVID-19 status. Additionally, participants required valid data on a minimum set of covariates, including age, sex and pre-pandemic mental health. Where possible, data within studies were weighted to be representative of their target population, accounting for sampling design, attrition up to the most recent pre-pandemic survey, and differential non-response to COVID-19 surveys. Weights were not available for GS or TwinsUK.

## Measures

Measures and derived variables are described below. Further detail is available in File S1, Table S2.

### Mental health outcomes

We assessed mental health (psychological distress, depression, anxiety, life satisfaction) using self-report measures across multiple time points of the pandemic. We standardised continuous scales within each study to permit comparability of estimates across studies. We also conducted analyses with dichotomous indicators using established cut-off scores for each scale.

### COVID-19

Self-reported COVID-19 was measured in each study and at each wave (see File S1). We used these measures to create a binary time-updated ‘ever had COVID-19’ (yes, no) variable.

We used information about time since COVID-19 to derive continuous and categorical time since infection variables in studies with available data (see Table S3-6). At each timepoint, for those who self-reported prior COVID-19, we created a continuous ‘duration since COVID-19 in weeks’ variable capturing the interval between estimated infection date and survey dates in which mental health outcomes were measured. We also created a variable with the following time since infection categories: no COVID-19; <4 weeks since infection; 4-12 weeks since infection; 12+ weeks since infection.

We derived two variables to examine differences between suspected and confirmed COVID-19. First, we created a categorical variable: no COVID-19, self-reported suspected COVID-19 (not test-confirmed), or self-reported test-confirmed COVID-19. Second, we created a variable using serology data from one timepoint (collected between April and June 2021) when available in the respective study, which was based on smaller subsamples of participants (see Table S6). We only used data from antibody tests with an immunoassay qualitative detection of antibodies against SARS-CoV-2 nucleocapsid (N) protein, of which a positive result (N-assay) is likely to identify SARS-CoV-2 infection. Using serology data combined with self-report information, we created a categorical variable: no COVID-19; self-report COVID-19 with negative serology; self-report COVID-19 with positive serology; and no self-reported COVID-19 with positive serology.

### Covariates

As available across studies, models were adjusted for the following covariates (see Table S7): sex (male; female); age (continuous); ethnicity (self-reported and coded into White or Non-White ethnic minorities); UK country of residence (England; Scotland; Wales; Northern Ireland); highest educational qualification (degree; no degree; parental education was used for the MCS cohort, who had not all completed their full-time education), pre-pandemic mental health (continuous), pre-pandemic chronic illness (yes; no), pre-pandemic disability (yes; no), pre-pandemic self-rated health (poor; fair or good), partnership status (partner; no partner), occupational classification (assessed through NS-SEC and coded into four categories: professional/managerial; intermediate; lower/manual; none/long term unemployed). All analyses were adjusted for data collection timepoints during the pandemic to control for overall population level changes in mental health at different stages of the pandemic.

## Analysis

### RQ1: Is COVID-19 illness associated with subsequent psychological distress, depression, anxiety and low life satisfaction?

We examined associations between ever-COVID-19 status and mental health using generalized estimation equations (GEE), specifying an unstructured correlation matrix, to account for correlations between repeated measures from the same individuals. For binary mental health outcomes, we used modified Poisson regression with robust standard errors to calculate relative risks.^22^ We ran models both unadjusted and fully adjusted for potential confounders.

### RQ2: Does the strength of the association vary according to time since infection?

First, we used GEE models to examine whether associations between COVID-19 and subsequent mental health varied according to time since infection categories: no COVID-19, <4 weeks, 4-12 weeks or 12+ weeks. Second, among those with COVID-19, we explored the relationship between continuous time since infection in weeks and mental health. We also ran models incorporating a quadratic term for continuous time since infection, to test for non-linearity.

### RQ3: Are associations modified by age, sex, ethnicity, education level, and pre-COVID-19 mental health?

We tested for interactions between COVID-19 and sex (male, female); ethnicity (White, Non-White ethnic minorities); highest educational qualification (degree, non-degree) and pre-pandemic mental health and life satisfaction (case, not case). We stratified analyses by age in age heterogeneous cohorts, using the following bands: 16-29, 30-49, 50-69, 70+ years.

### RQ4: Do associations between COVID-19 and subsequent mental health differ between suspected vs confirmed infection?

First, using GEE models, we examined whether associations between COVID-19 and mental health differed between those with self-reported suspected COVID-19 vs test-confirmed COVID-19. Second, we explored differences in association for those with self-report suspected vs serology-detected SARS-CoV-2. Given the timing of serology assessments - in many cohorts alongside or after the most recent mental health assessment and not time varying – we examined associations with mental health at the most recent time point only (linear and modified Poisson regression) for those who: did not have COVID-19 (reference group); self-reported COVID-19 and had serology evidence of SARS-CoV-2; self-reported COVID-19 but no serology-detected SARS-CoV-2; did not self-report COVID-19 but had evidence of SARS-CoV-2 infection. In an exploratory analysis, we also tested the association between SARS-CoV-2 serology status (positive VS negative) with most recent mental health scores.

### Sensitivity analyses

In NSHD, NCDS, BCS70, NS and MCS, participants who reported “unsure” as to whether they had COVID-19 were grouped as COVID-19 cases if they reported an estimated infection date, or as non-cases if they did not report a date. In sensitivity analyses, we compared these results to findings in which those reporting ‘unsure’ were: a) categorised as not having had COVID-19 and b) retained as a separate category.

### Pooled estimates

We pooled estimates across studies using random effects meta-analysis with restricted maximum likelihood. Within age heterogeneous cohorts, we pooled estimates from age-stratified analyses using the following age bands: 16-29, 30-49, 50-69, 70+, and age homogenous cohorts were grouped within the appropriate age band. We reported heterogeneity indices using the I^2^, and where possible also T^2^ and 95% prediction intervals (95% PI).^23^

## Results

We conducted analyses using 11 different longitudinal studies (*k*), with 54,442 participants in total. Individual study sample sizes ranged from 1,721 in NSHD to 14,154 in USoc. Descriptive statistics for exposures, outcomes and covariates are presented in Tables S4-6 and S8-9.

### Descriptive analysis

By the first survey timepoint (April – June 2020), across all studies between 5.4% (NSHD) and 19.3% (NS) of participants self-reported COVID-19. By the final timepoint for each study (Nov 2020 - April 2021) between 11.1% (NSHD) and 45.1% (MCS) self-reported COVID-19. Serology data indicated that between 4.7% (NSHD) and 22.7% (MCS) had positive antibody results indicating natural SARS-CoV-2 infection. Of those with information on COVID-19 and serology data, between 2.6% (NSHD) and 18.1% (MCS) had both self-reported and serology-confirmed COVID-19, whereas those with self-reported COVID-19 but negative serology data ranged 8.5% (NSHD) to 31.7% (MCS). The proportion of people with positive serology data who did not self-report COVID-19 ranged from 2.1% (NSHD) to 17.3% (TwinsUK).

### RQ1 Associations between COVID-19 and subsequent mental health

Unadjusted results can be found in Figure S1. Pooled adjusted estimates from meta-analyses indicated that COVID-19 was associated with an increase in subsequent psychological distress (standardized difference in outcome between those with and without self-reported COVID-19) = 0.10 [95%CI: 0.06; 0.13], I^2^ = 42.8%; *k* = 8), depression (0.08 [0.05; 0.10], I^2^ = 20.8%; *k* = 9), and anxiety (0.08 [0.05; 0.10], I^2^ = 0.0; *k* = 9), and negatively associated with life satisfaction (−0.06 [-0.08; -0.04], I^2^ = 29.2%; *k* = 10). Results were consistent for binary outcomes in terms of effect size and direction (psychological distress RR = 1.15 [95%CI: 1.05; 1.25], I^2^ = 88.8%; *k* = 8) (See Figure 1b and Figure S2). Meta-analysed coefficients for all research questions are reported in Tables S10 and S11.

**Figure 1a (left) & 1b (right):**
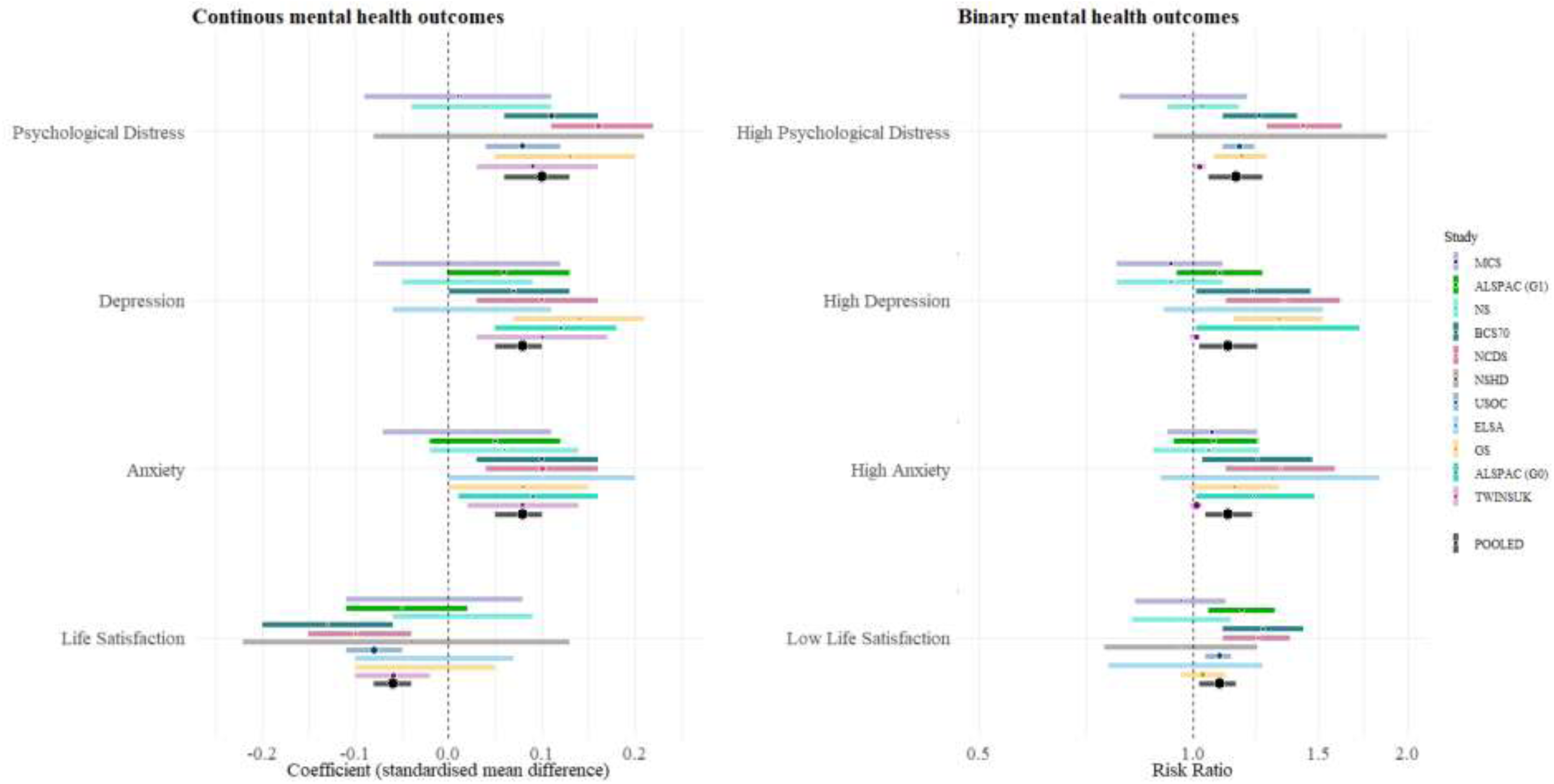
Estimates from the longitudinal GEE models with ever-COVID-19 exposure and mental health outcomes (RQ1) for each included study and the overall pooled estimate using random-effects meta-analysis.

### RQ2 Associations between COVID-19 and mental health by time since infection

We examined time since infection using both categorical and continuous variables. Pooled results indicated that the association between COVID-19 and mental health did not differ according to time since infection for psychological distress, with similar associations across duration categories, although heterogeneity increased with time since infection: 4 weeks (0.10 [0.04; 0.16], I^2^ = 0.0%; *k* = 5), 4-12 weeks (0.10 [0.04; 0.17], I^2^ = 21.6%; *k* = 5), and 12+ weeks (0.10 [0.04; 0.15], I^2^ = 62.8%; *k* = 5). Heterogeneity between time since COVID-19 infection categories was I^2^=18.1%. This pattern of results was consistent for depression, anxiety and life satisfaction (Figure 2 and Figures S3-6). For those with COVID-19, no association was found between continuous time since infection in weeks and all outcomes. We examined non-linearity with a quadratic term and found no evidence of a non-linear association.

**Figure 2:**
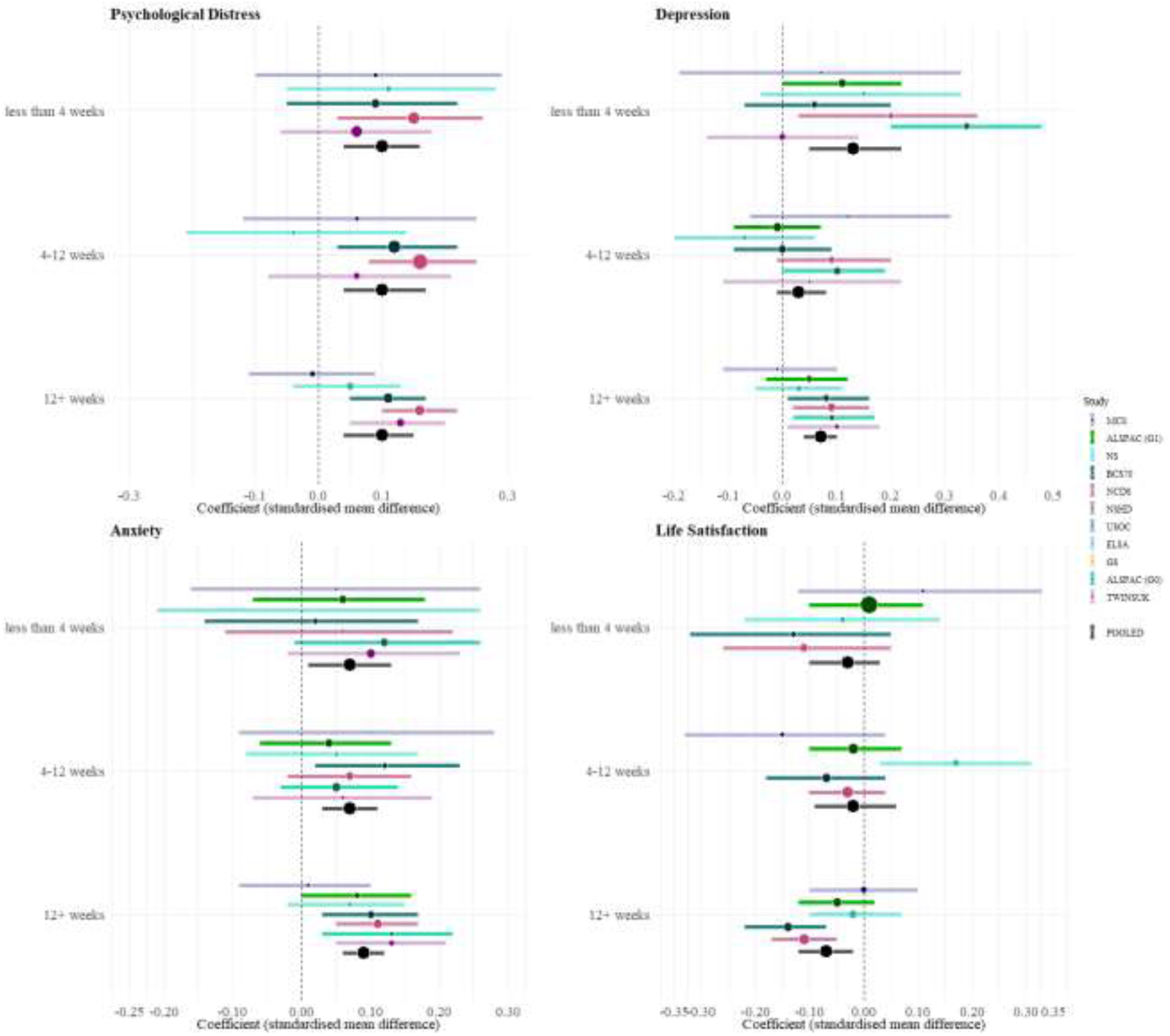
Time since infection (in categories) and continuous mental health outcomes (RQ2) for each included study and the overall pooled estimate.

### RQ3 Subgroup differences by age, sex, ethnicity, education level, and pre-pandemic mental health

Stratified analyses indicated that associations between COVID-19 and mental health were present in all age groups, with some evidence of stronger effects in middle-aged and older groups, for example, among those aged 50-69 years: psychological distress (0.13 [0.10; 0.15], I^2^ = 0%; *k* = 5), depression (0.10 [0.06; 0.15], I^2^ = 44.2%; *k* = 6), and anxiety (0.10 [0.06; 0.13], I^2^ = 0.0%; *k* = 6), and lower life satisfaction (−0.07 [-0.11; -0.04], I^2^ *=* 30.1%; *k* = 5). This pattern was similar for those aged 70 years and older (Figure 3). We did not find evidence of interaction between COVID-19 and: sex, education, ethnicity or pre-pandemic mental health (see Figures S7-10).

**Figure 3:**
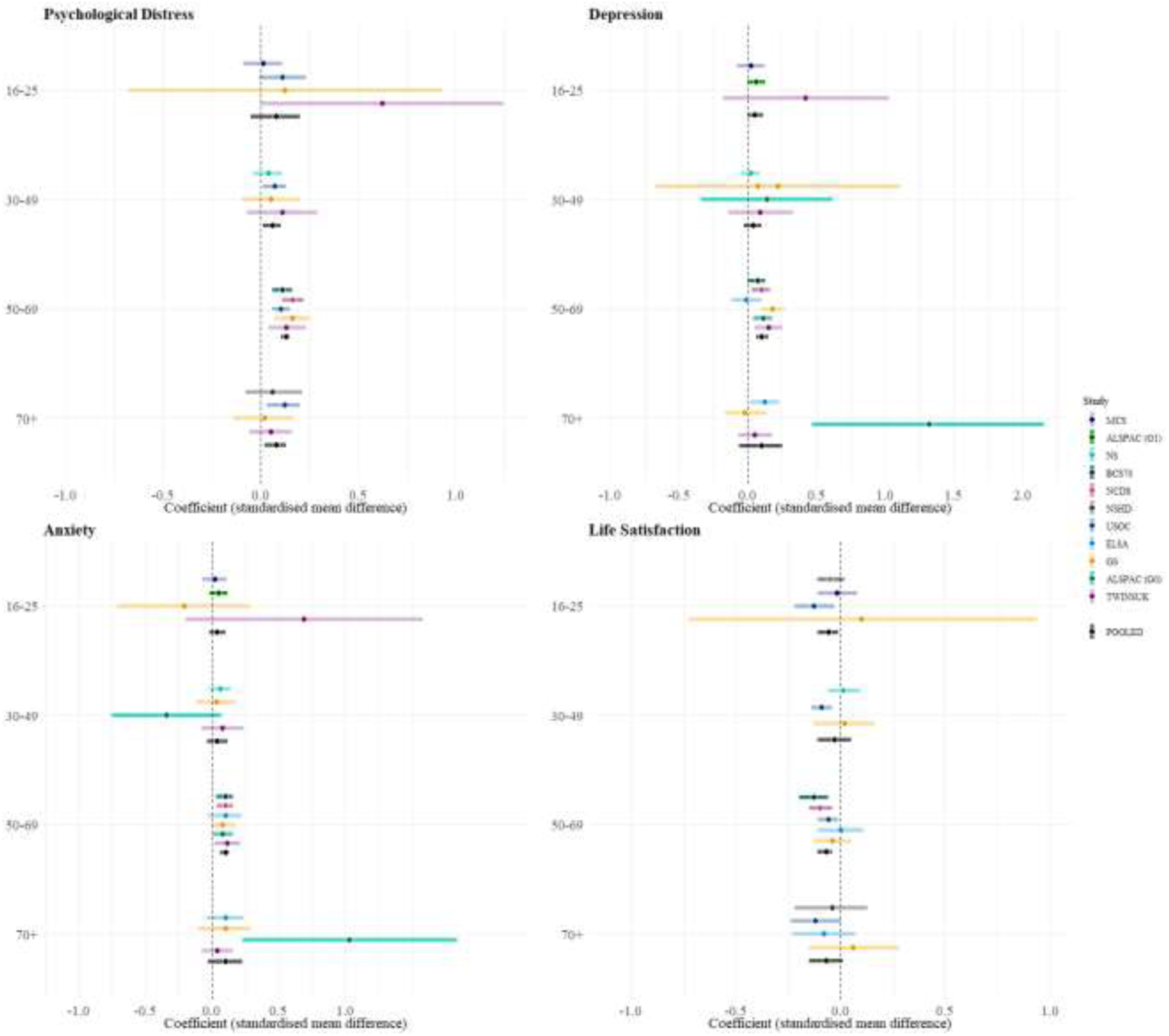
Age stratified estimates for the ever-COVID-19 exposure and mental health outcomes (RQ3) for each included study and the overall pooled estimate. Notes.

### RQ4 Associations between COVID-19 and mental health for suspected COVID-19 relative to: A) test-confirmed COVID-19 and B) serology-confirmed COVID-19

First, we examined whether associations differed between those with suspected COVID-19 vs test-confirmed COVID-19, both based on self-report. *Suspected* (0.09 [0.07; 0.11], I^2^ = 0.0%; *k* = 8) and *test-confirmed* COVID-19 (0.11 [0.02; 0.19], I^2^ = 68.3%; *k* = 8) were associated with increased psychological distress, with similar patterns for depression and anxiety, although only *suspected* COVID-19 was associated with lower life satisfaction (Figure 4 and Figures S11-14).

**Figure 4:**
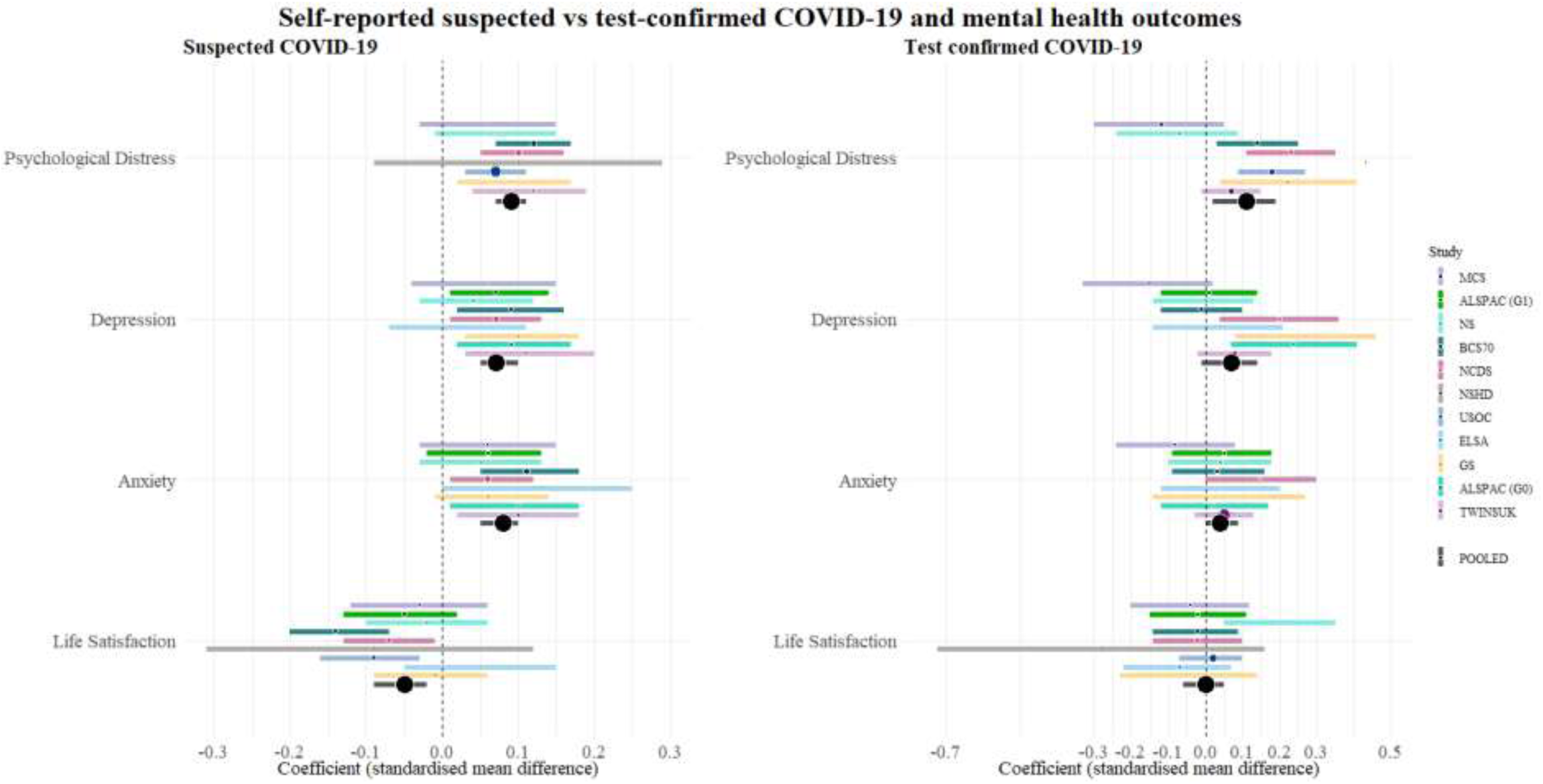
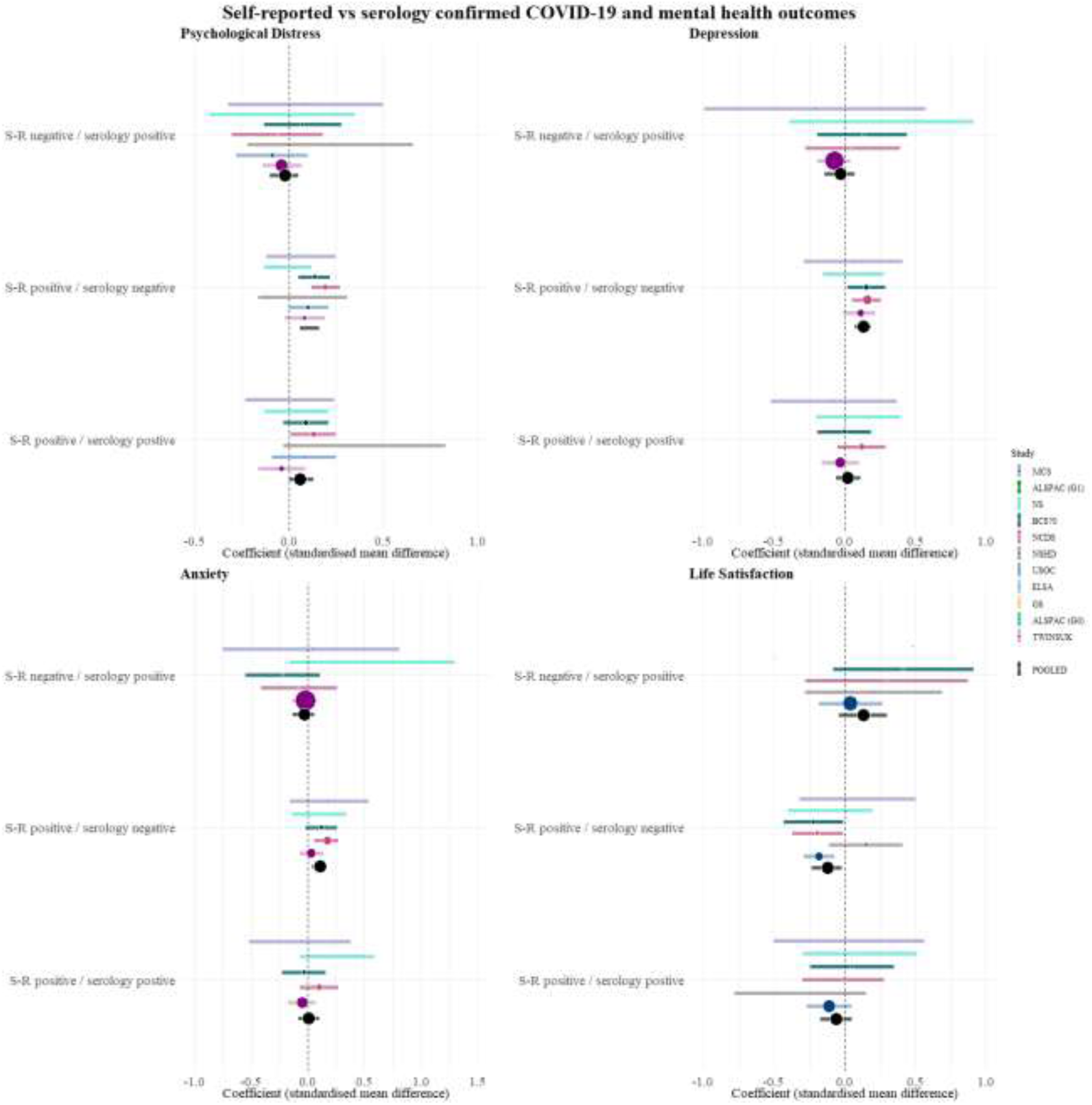
Suspected vs. Test-confirmed COVID-19 infection and mental health outcomes (RQ4)

Second, we examined whether associations varied based on combined self-report and serology data (Table S6). Those who self-reported COVID-19 but had negative serology had higher levels of psychological distress (0.11 [0.06; 0.16], I^2^ = 29.5%; *k* = 7), depression, anxiety and lower life satisfaction than those without COVID-19 based on self-report and serology. Similar patterns were found for psychological distress and life satisfaction in those with self-reported COVID-19 and positive serology, but associations were not found for depression or anxiety (Figure 4). For those who were serology positive but did not self-report COVID-19, we did not find associations with mental health outcomes (psychological distress -0.02 [−0.09; 0.06], I^2^ = 0.0%; *k* = 7; see Figure 4 and Figures S15-18). In an additional exploratory analysis comparing those with positive serology to those with negative serology, we did not find evidence of differences in psychological distress (0.02 [-0.03; 0.07], I^2^ = 12.8%; *k* = 7) or other mental health outcomes (Figure S19).

### Sensitivity analyses

To examine the potential impact of the ‘unsure’ categorisation in MCS, NS, BCS, NCDS and NSHD, we conducted a sensitivity analysis for RQ1, grouping those reporting ‘unsure’ with those who did not report having had COVID-19. Associations remained consistent (psychological distress = 0.09 [95%CI: 0.05; 0.12, I^2^ = 51.9%]); Figure S20-21). In the second sensitivity analysis, both self-reported COVID-19 (psychological distress = 0.10 [95%CI: 0.00; 0.19, I^2^ = 71%] and unsure COVID-19 (psychological distress = 0.10 [95%CI: 0.04; 0.15, I^2^ = 43.8%] showed associations with poorer mental health (Figures S22-25).

## Discussion

Our findings indicate that COVID-19 illness was associated with deterioration in mental health outcomes in the UK population. We did not find evidence of change in this association over time during the first few months’ post-infection. Subgroup analysis indicated no differences by sex, ethnicity, education or pre-pandemic mental health; whereas associations were stronger in older age groups. Lastly, we observed similar associations for both suspected and confirmed COVID-19, suggesting that the associations could relate to experience of disease (rather than exposure to the virus per se), highlighting the salience of psychosocial mechanisms. Taken together, the limited attenuation in association over time since infection and findings involving serology-confirmed infection raise the possibility that the effects observed might not be specific to COVID-19 but could still reflect the mental health impact of illness during this period and/or be explained by other factors.

Our findings demonstrate associations between COVID-19 and deterioration in mental health while controlling for overall effects of timing throughout the first year of the pandemic, adding to existing evidence which has been mixed to-date.^3,4^ The effects observed (6-10% change of a standard deviation for outcomes on a continuous scale and 9-15% increased risk of clinical caseness), has substantial implications when considered at the population level, especially given high infection rates.

We did not observe improvement in mental health in the immediate months post-infection.^3^ Studies with longer term follow-up examining recovery in symptoms are needed to assess the duration of symptoms experienced post-infection.

We found that COVID-19 was associated with poorer mental health in all age groups, with some evidence of stronger associations for people aged 50 years and older. This might reflect that older people are more likely to experience more severe COVID-19 and potentially also greater worry around infection due to their age and higher likelihood of pre-existing health conditions. These findings could also reflect increased risk of microvascular or neurological changes post COVID-19, which has been associated with depression and anxiety phenotypes in older adults.^24–26^ On the other hand, we found no differences by sex, ethnicity, education level and previous mental health problems. Previous studies have shown that overall mental health impacts of the pandemic have been greatest in adults aged 25-44 years, women, and those with higher degrees;^19^ suggesting that mechanisms through which COVID-19 illness impacts mental health may differ from those underpinning wider effects of the pandemic.

Our analyses benefited from the use of serology data in addition to information on self-reported COVID-19.^26^ When comparing associations for subgroups based on self-report and serology status, self-reported COVID-19 illness combined with either negative or positive serology was associated with poorer mental health, whereas no association was found for positive serology without self-reported COVID-19. Similarly, in an additional exploratory analysis, we did not find evidence of differences in mental health outcomes for those with positive and negative serology. Various potential mechanisms have been posited to underlie associations between COVID-19 and psychological distress, including systemic inflammation and changes in the brain associated with COVID-19,^27^ and psychosocial mechanisms including social isolation and, worries about possible outcomes and infecting others. One possibility for our findings is that contextual and psychosocial aspects of COVID-19, such as feeling unwell and worrying about potential health, social and economic consequences, could be stronger predictors of poor mental health outcomes than any specific neurological consequences of SARS-CoV-2 infection. The lack of association with mental health when serology data detected SARS-CoV-2 infections that had not been reported, i.e., cases where participants were unaware of the infection, also support this conclusion. However, this group were likely to have had mild or asymptomatic COVID-19. In addition, it is possible that those who were particularly concerned about consequences of infection were more likely to report perceived infection. It should also be noted that only a subset of studies and samples had serology data, thereby substantially reducing power in these analyses. Additionally, only one timepoint of serology assessment was completed after the most recent self-report data thereby limiting the conclusions that can be drawn. Further, antibody levels following SARS-CoV-2 infection have been found to wane over time,^27^ which could have led to misclassification based on serology data.

The use of multiple longitudinal studies is a significant strength and allowed us to control for time period effects and important pre-pandemic factors including mental health, physical health and socio-economic factors. Thus, in comparison to previous studies using electronic health records or pandemic-specific online surveys, we were able to include more detailed information on a wider range of pre-pandemic variables. Rich antecedent data in longitudinal prospective studies also allowed most studies to be weighted for non-response (reducing potential bias from selection into analysed samples). However, while we were able to control for important confounders, we cannot definitively attribute changes in mental health to COVID-19 illness. Included studies used varying measures to assess COVID-19 and mental health outcomes. These were carefully reviewed and harmonised across studies, and in meta-analysis the heterogeneity of estimates between studies was small for most outcomes. Nonetheless, measurement error of the exposure is a potential limitation, given that our main exposure variables were based on self-reported COVID-19; although the additional analysis including serology data helps to mitigate against this. Previous studies have mainly focused on severe COVID-19 or severe mental illness.^8,10^ Our results add to existing evidence by capturing broader and subclinical mental health impacts of COVID-19 illness in the population. However, further possible limitations are the lack of data available to examine possible variation in associations by COVID-19 severity, that only infections in the first year of the pandemic have been assessed, and that longitudinal follow-up is limited at present.

### Implications and conclusions

Our findings suggest that people who self-reported COVID-19 in the first year of the pandemic were subsequently more likely to experience poorer mental health outcomes. Our findings involving serology-confirmed infection, and the lack of attenuation in association over time, suggest that these associations might not be specific to SARS-CoV-2 infection and potentially reflect consequences of feeling unwell, anxieties related to a novel infection and infecting others or other factors such as social isolation and loss of pay. Further research is needed to investigate these possible underlying mechanisms and to examine whether associations persist over longer follow-up periods. Our findings highlight the important population mental health consequences of infection and disease itself separately from the potential impacts of the pandemic more widely (e.g., infection control measures). Given the high prevalence of COVID-19 in the UK and worldwide, these findings have important public health implications, highlighting the need for greater post-infection mental health support in both clinical and community settings.

## Supporting information

Supplementary material Variable coding

## Data Availability

All data used in this research can either be access online or be requested from the participating organizations. More information for each dataset is provided via the following links:
MCS,NS, BCS70 and NCDS COVID Data: http://doi.org/10.5255/UKDA-SN-8658-3 
MCS: http://doi.org/10.5255/UKDA-SN-8682-2
NS:  http://doi.org/10.5255/UKDA-SN-5545-8 
BCS70: http://doi.org/10.5255/UKDA-SN-8547-1 
NCDS: http://doi.org/10.5255/UKDA-SN-7669-1
NSHD: https://www.nshd.mrc.ac.uk/
TWINS: https://twinsuk.ac.uk/resources-for-researchers/our-data/
ALSPAC-G1 & ALSPAC-G0  http://www.bristol.ac.uk/alspac/researchers/our-data/
USOC http://doi.org/10.5255/UKDA-SN-8644-10 
ELSA http://doi.org/10.5255/UKDA-SN-5050-23
GS https://www.ed.ac.uk/generation-scotland/for-researchers/covidlife 
https://www.ed.ac.uk/generation-scotland/for-researchers/generation-scotland 

## Author contributions

Conceptualisation: PP, EJT, JS, BM, CS, NC, VK, ASFK

Methodology: EJT, JS, BM, PP, RS, KT, MP, NC, VK, PZ, RS, ASFK, CH, MJG, JM, KM

Data Analysis: EJT, JS, BM, CH, ASFK, RS, PZ

Visualisation: JS, EJT

Writing- drafting: EJT, JS, BM, PP

Writing- review: all authors

Supervision: PP, CS, NC

Data curation: EJT, JS, BM, CH, ASFK, RS, PZ, EM, JP, PK

Funding: NC, PP, GP, VK, CS, NT, DJP, GBP

## Funding Acknowledgements

This work was supported by the National Core Studies, an initiative funded by UKRI, NIHR and the Health and Safety Executive. The COVID-19 Longitudinal Health and Wellbeing National Core Study was funded by the Medical Research Council (MC_PC_20059) and the CONVALESCENCE study was funded by the NIHR (CONVALESCENCE grant COV-LT-0009). Full funding acknowledgements for each individual study can be found as part of Supplementary File 1.

