## Supplementary material for "Mental health outcomes following COVID-19 infection: Evidence from 11 UK longitudinal population studies"

**Supplementary Materials**

**Supplementary Tables**

### Table S1. Ethics and data access statements for each study

| **NSHD, NCDS, BCS70**, **NS** and **MCS** | The most recent sweeps of the **NSHD, NCDS, BCS70**, **Next Steps** and **MCS** have all been granted ethical approval by the National Health Service (NHS) Research Ethics Committee and all participants have given informed consent. Data for NCDS (SN 6137), BCS70 (SN 8547), Next Steps (SN 5545), MCS (SN 8682) and all four COVID-19 surveys (SN 8658) are available through the UK Data Service. NSHD data are available on request to the NSHD Data Sharing Committee. Interested researchers can apply to access the NSHD data via a standard application procedure. Data requests should be submitted to; further details can be found at <http://www.nshd.mrc.ac.uk/data.aspx>. doi:10.5522/NSHD/Q101; doi:10.5522/NSHD/Q10. |
| --- | --- |
| **ALSPAC G0** and **ALSPAC G1** | Ethical approval was obtained from the **ALSPAC** Ethics and Law Committee and the Local Research Ethics Committees. The study website contains details of all the data that is available through a fully searchable data dictionary and variable search tool: <http://www.bristol.ac.uk/alspac/researchers/our-data>. ALSPAC data is available to researchers through an online proposal system. Information regarding access can be found on the ALSPAC website (<http://www.bristol.ac.uk/media-library/sites/alspac/documents/researchers/data-access/ALSPAC_Access_Policy.pdf>). Study data were collected and managed using REDCap electronic data capture tools hosted at the University of Bristol. [REDCap](https://www.project-redcap.org/) (Research Electronic Data Capture) is a secure, web-based software platform designed to support data capture for research studies. |
| **USOC** | The University of Essex Ethics Committee has approved all data collection for the **Understanding Society** main study and COVID-19 waves. No additional ethical approval was necessary for this secondary data analysis. All data are available through the UK Data Service (SN 6614 and SN 8644). |
| **ELSA** | Waves 1-9 of **ELSA** were approved through the National Research Ethics Service, while the COVID-19 Sub-study was approved by the UCL Research Ethics Committee. All participants provided informed consent. All data are available through the UK Data Service (SN 8688 and 5050). |
| **GS** | **Generation Scotland** obtained ethical approval from the East of Scotland Committee on Medical Research Ethics (on behalf of the National Health Service). Reference number 20/ES/0021. Access to data is approved by the Generation Scotland Access Committee. See <https://www.ed.ac.uk/generation-scotland/for-researchers/access> or for further details. |
| **TWINSUK** | All wave of **TwinsUK** have received ethical approval associated with TwinsUK Biobank (19/NW/0187), TwinsUK (EC04/015) or Healthy Ageing Twin Study (H.A.T.S) (07/H0802/84) studies from NHS Research Ethics Committees at the Department of Twin Research and Genetic Epidemiology, King’s College London. The TwinsUK Resource Executive Committee (TREC) oversees management, data sharing and collaborations involving the TwinsUK registry (for further details see <https://twinsuk.ac.uk/resources-for-researchers/access-our-data/>). |

### Table S2. Details of mental health measures in each longitudinal study

| **Study** | **Details of mental health measures** |
| --- | --- |
| **MCS** | The K-6 is a 6-item measure of psychological distress (i.e., general anxiety and depression). Responses are rated on a 5-point Likert-type scale, and capture distress over a period of four weeks prior to administration of the scale. Scores range from 0 to 24, with a conservative cut-off of 13+ applied to indicate probable psychological distress. |
| **ALSPAC** | Self-reported depressive symptoms were measured using the short mood and feelings questionnaire (SMFQ). The SMFQ is a 13-item questionnaire that measures the presence of depression symptoms in the previous two weeks and was administered via postal questionnaire or in research clinics. Each item is scored between 0-2, resulting in a summed score between 0-26. Depression severity can be rated in the following score bands: 0-4 none, 5-9 mild, 10-14 moderate, 15-19 moderately severe, 20-27 severe. ALSPAC also used the seven-item Generalized Anxiety Disorder Scale (GAD-7), which is a validated, self-report measure of anxiety used widely by healthcare professionals. Individuals were asked to consider how frequently they have been bothered by a number of problems in the previous two weeks and respond on a four-point Likert scale from 0 “Not at all” to 3 “Nearly every day”. The cut-off points for mild, moderate and severe anxiety, are 5, 10, and 15 respectively, and the maximum score is 21. |
| **NS** | The 12-item General Health Questionnaire (GHQ) was used to detect symptoms of psychological distress in NS and USoc. The GHQ is a screening instrument designed to detect symptoms of psychological distress (i.e. general anxiety and depression). Each item is scored 0-3 resulting in scores ranging from 0-36. There is an alternative scoring where each item is scored as 0-0-1-1. |
| **BCS70, NCDS** | The 9-item version of the Malaise Inventory was used to assess general psychological distress. Items are scored using a simple ‘Yes/No’ response, meaning continuous scores range from 0-9. Scores of four or more are indicative of probable psychiatric distress. |
| **NSHD** | The General Health Questionnaire (GHQ) was also used to detect symptoms of psychological distress in NSHD. For the pre-pandemic sweep, the 28-item GHQ was used, though the 12-item GHQ was used for the subsequent 3 sweeps. Each item is scored as 0-0-1-1, resulting in scores ranging from 0-12 with higher scores indicating greater likelihood of mental ill health, and a threshold of 4 was used for binary analyses. |
| **USoc** | The 12-item General Health Questionnaire (GHQ) was used, as described for NS. |
| **ELSA** | Depressive symptoms were measured using an abbreviated 8-item version of the validated Center for Epidemiologic Studies Depression Scale (CES-D). Respondents were asked whether they had experienced any depressive symptoms, such as feeling sad or having restless sleep, in the week prior to interview. For the binary classification, we considered respondents who reported four or more depressive symptoms on the CES-D scale as having elevated depressive symptoms. |
| **GS** | Depression and anxiety were assessed during the pandemic sweeps using the GAD-7 and the Patient Health Questionnaire (PHQ-9). The PHQ-9 is a nine-item, validated tool for the assessment of depressive symptoms experienced in the previous two weeks. Participants are asked to indicate how often they have been bothered by problems such as “Little interest or pleasure in doing things?” on a four-point Likert scale from 0 “Not at all” to 3 “Nearly every day”. The score is the sum of the nine items, to a total of 27. A score of 10 or more indicates major depression. The 28-item GHQ was used to assess pre-pandemic psychological distress, and so a comparable composite measure was created from the GAD-7 and the PHQ-9 scales to enable evaluation of change over time. Correlations between each GHQ-28 item and items of the GAD-7 and PHQ-9 scales were examined, and the text of the most highly correlated (r > 0.25) questions was then reviewed and sense-checked; matched items are presented in the table below. Cut-off scores were determined using ROC curves, which found a general cut-off of >4, a depression cut-off of >5 and an anxiety cut-off of >2. The latter matched the general “proportion” of PHQ-9/GAD-7 scores required for a cut-off. |
| **TwinsUK** | The Hospital Anxiety and Depression Scale (HADS) is a 14-item scale used to measure levels of psychiatric distress in non-psychiatric patient populations. Responses are indicated on a 4-point ordinal Likert scale. |

Further information on specific items, response options and coding for all involved variables in this study can be found here: https://osf.io/x2mu5/

### Table S3. Descriptives for reporting of COVID-19 infection in each wave of data collection and % for the ever-COVID-19 exposure variable per wave of each included study

| Study | Wave Timing (month-year) | N per wave | N reporting C-19 (per wave) | % Time updated ever- COVID-19 |
| --- | --- | --- | --- | --- |
| MCS | May-20 | 2350 | 320 | 13.6 |
| MCS | Sept-Oct 2020 | 2807 | 954 | 34.2 |
| MCS | Feb-March 2021 | 3837 | 1523 | 45.1 |
| ALSPAC-G1 | April 2020 | 2011 | 302 | 15.0 |
| ALSPAC-G1 | May-20 | 2261 | 441 | 19.5 |
| ALSPAC-G1 | December 2020 | 2777 | 647 | 23.3 |
| NS | May-20 | 1678 | 324 | 19.3 |
| NS | Sept-Oct 2020 | 3093 | 946 | 30.9 |
| NS | Feb-March 2021 | 3482 | 1171 | 39.8 |
| BCS70 | May-20 | 3,436 | 661 | 19.2 |
| BCS70 | Sept-Oct 2020 | 4253 | 1161 | 27.8 |
| BCS70 | Feb-March 2021 | 4629 | 1305 | 34.6 |
| NCDS | May-20 | 4456 | 637 | 14.3 |
| NCDS | Sept-Oct 2020 | 5345 | 1074 | 20.5 |
| NCDS | Feb-March 2021 | 5784 | 1184 | 26.0 |
| NSHD | May-June 2020 | 1432 | 87 | 5.4 |
| NSHD | Sept-Oct 2020 | 1677 | 145 | 7.6 |
| NSHD | Feb-March 2021 | 1536 | 173 | 11.1 |
| USoC | Apr-20 | 12,871 | 1,539 | 11.5 |
| USoC | May-20 | 11,642 | 1,502 | 12.3 |
| USoC | Jun-20 | 11,189 | 1,499 | 12.7 |
| USoC | Jul-20 | 10,944 | 1,551 | 13.4 |
| USoC | Sep-20 | 10,332 | 1,631 | 15.4 |
| USoC | Nov-20 | 9,742 | 1,756 | 17.7 |
| USoC | Jan-21 | 9,646 | 2,109 | 22.1 |
| USoC | Mar-21 | 10,150 | 2,395 | 23.8 |
| ELSA | June-July 2020 | 4710 | 402 | 9.9 |
| ELSA | Nov-Dec 2020 | 4889 | 129 | 12.4 |
| GS | Apr-20 | 3937 | 314 | 8.0 |
| GS | Jul-20 | 2924 | 421 | 10.7 |
| GS | Feb-21 | 2728 | 540 | 13.7 |
| ALSPAC-G0 | April 2020 | 2774 | 342 | 12.3 |
| ALSPAC-G0 | May-20 | 2997 | 431 | 14.4 |
| ALSPAC-G0 | December 2020 | 3483 | 585 | 16.7 |
| TwinsUK | Jul-20 | 2437 | 293 | 9.3 |
| TwinsUK | Oct-20 | 2291 | 308 | 15.9 |
| TwinsUK | Apr-21 | 1932 | 294 | 19 |

### Table S4. Counts and percentages for those self-reporting suspected and confirmed COVID-19 per wave of each included study

| **Cohort** | **Wave Timing** | **Total N** | **N (%) reporting no C-19** | | **N (%) reporting C-19 suspected** | | **N (%) reporting C-19 confirmed** | | **N (%) reporting unsure about whether have had COVID-19** |
| --- | --- | --- | --- | --- | --- | --- | --- | --- | --- |
| MCS_CM | May-20 | 2350 | 1710 | (72.77) | 143 | (6.09) | 6 | (0.26) | 491 |
| MCS_CM | Sept-Oct 2020 | 2807 | 1853 | (66.01) | 301 | (10.72) | 50 | (1.78) | 603 |
| MCS_CM | Feb-March 2021 | 3837 | 2314 | (60.31) | 461 | (12.01) | 500 | (13.03) | 562 |
| MCS_CM (unsure recoded) | May-20 | 2350 | 2030 | (86.38) | 314 | (13.36) | 6 | (0.26) |  |
| MCS_CM (unsure recoded) | Sept-Oct 2020 | 2807 | 1853 | (66.01) | 904 | (32.21) | 50 | (1.78) |  |
| MCS_CM (unsure recoded) | Feb-March 2021 | 3837 | 2314 | (60.31) | 1023 | (26.66) | 500 | (13.03) |  |
| ALSPAC-G1 | April 2020 | 2011 | 1709 | (85) | 264 | (13.1) | 38 | (1.9) |  |
| ALSPAC-G1 | May-20 | 2261 | 1820 | (80.5) | 395 | (17.5) | 46 | (2) |  |
| ALSPAC-G1 | December 2020 | 2777 | 2130 | (76.7) | 499 | (18) | 148 | (5.3) |  |
| NS | May-20 | 1678 | 1125 | (67.04) | 164 | (9.77) | 12 | (0.72) | 377 |
| NS | Sept-Oct 2020 | 3093 | 2147 | (69.41) | 318 | (10.28) | 67 | (2.17) | 561 |
| NS | Feb-March 2021 | 3482 | 2311 | (66.37) | 371 | (10.65) | 331 | (9.51) | 469 |
| NS (unsure recoded) | May-20 | 1678 | 1354 | (80.69) | 312 | (18.59) | 12 | (0.72) |  |
| NS (unsure recoded) | Sept-Oct 2020 | 3093 | 2147 | (69.41) | 879 | (28.42) | 67 | (2.17) |  |
| NS (unsure recoded) | Feb-March 2021 | 3482 | 2311 | (66.37) | 840 | (24.12) | 331 | (9.51) |  |
| BCS70 | May-20 | 3436 | 2342 | (68.16) | 291 | (8.47) | 13 | (0.38) | 790 |
| BCS70 | Sept-Oct 2020 | 4253 | 3092 | (72.7) | 380 | (8.93) | 63 | (1.48) | 718 |
| BCS70 | Feb-March 2021 | 4629 | 3280 | (70.86) | 415 | (8.97) | 317 | (6.85) | 617 |
| BCS70 (unsure recoded) | May-20 | 3436 | 2775 | (80.76) | 648 | (18.86) | 13 | (0.38) |  |
| BCS70 (unsure recoded) | Sept-Oct 2020 | 4253 | 3092 | (72.7) | 1098 | (25.82) | 63 | (1.48) |  |
| BCS70 (unsure recoded) | Feb-March 2021 | 4629 | 3324 | (71.81) | 988 | (21.34) | 317 | (6.85) |  |
| NCDS | May-20 | 4456 | 3353 | (75.25) | 241 | (5.41) | 17 | (0.38) | 845 |
| NCDS | Sept-Oct 2020 | 5345 | 4271 | (79.91) | 330 | (6.17) | 47 | (0.88) | 697 |
| NCDS | Feb-March 2021 | 5784 | 4600 | (79.53) | 342 | (5.91) | 258 | (4.46) | 584 |
| NCDS (unsure recoded) | May-20 | 4456 | 3819 | (85.7) | 620 | (13.91) | 17 | (0.38) |  |
| NCDS (unsure recoded) | Sept-Oct 2020 | 5345 | 4271 | (79.91) | 1027 | (19.21) | 47 | (0.88) |  |
| NSHD | May-June 2020 | 1432 | 1278 | (89.33) | 27 | (1.64) | 3 | (0.1) | 124 |
| NSHD | Sept-Oct 2020 | 1677 | 1532 | (92.00) | 43 | (2.58) | 10 | (0.38) | 92 |
| NSHD | Feb-March 2021 | 1536 | 1396 | (87.87) | 37 | (3.45) | 26 | (1.29) | 77 |
| NSHD (unsure recoded) | May-June 2020 | 1432 | 1345 | (94.57) | 84 | (5.32) | 3 | (0.1) |  |
| NSHD (unsure recoded) | Sept-Oct 2020 | 1677 | 1540 | (92.77) | 127 | (6.85) | 10 | (0.38) |  |
| NSHD (unsure recoded) | Feb-March 2021 | 1536 | 1402 | (91.45) | 108 | (7.26) | 26 | (1.29) |  |
| USoC | Apr-20 | 12,871 | 11,375 | (88.80) | 1,477 | (11.00) | 19 | (0.15) |  |
| USoC | May-20 | 11,642 | 11,347 | (97.55) | 281 | (2.28) | 14 | (0.17) |  |
| USoC | Jun-20 | 11,189 | 11,028 | (98.62) | 140 | (1.25) | 21 | (0.14) |  |
| USoC | Jul-20 | 10,944 | 10,817 | (98.80) | 118 | (1.10) | 9 | (0.07) |  |
| USoC | Sep-20 | 10,332 | 10,178 | (98.33) | 136 | (1.50) | 18 | (0.16) |  |
| USoC | Nov-20 | 9,742 | 9,475 | (97.20) | 130 | (1.42) | 137 | (1.38) |  |
| USoC | Jan-21 | 9,646 | 9,214 | (95.08) | 141 | (0.02) | 291 | (3.42) |  |
| ELSA* | June-July 2020 | 4710 | 4308 | (90.14) | 375 | (9.15) | 27 | (0.68) |  |
| ELSA | Nov-Dec 2020 | 4889 | 4760 | (96.22) | 11 | (0.3) | 118 | (3.47) |  |
| GS | Apr-20 | 3937 | 3622 | (92.00) | 305 | (7.75) | 9 | (0.23) |  |
| GS | Jul-20 | 2924 | 2652 | (90.70) | 260 | (8.89) | 12 | (0.41) |  |
| GS | Feb-21 | 2728 | 2420 | (88.71) | 232 | (8.50) | 74 | (2.71) |  |
| GS | Total | 3937 | 3397 | (86.28) | 460 | (11.68) | 80 | (2.03) |  |
| ALSPAC-G0 | April 2020 | 2774 | 2432 | (87.7) | 319 | (11.5) | 23 | (0.8) |  |
| ALSPAC-G0 | May-20 | 2997 | 2566 | (85.6) | 407 | (13.6) | 24 | (0.8) |  |
| ALSPAC-G0 | December 2020 | 3483 | 2899 | (83.2) | 498 | (14.3) | 87 | (2.5) |  |
| TwinsUK | Jul-20 | 2766 | 2473 | (89.4) | 270 | (9.8) | 23 | 0.8 |  |
| TwinsUK | Oct-20 | 2299 | 1991 | (86.6) | 163 | (7.1) | 145 | 6.3) |  |
| TwinsUK | Apr-21 | 1968 | 1709 | (86.8) | 166 | (8.4) | 93 | 4.7) |  |
| TwinsUK | Total | 3322 | 2688 | (81) | 325 | (9.8) | 307 | 9.2) |  |

*Weighted

### Table S5. Descriptive statistics for self-reported COVID-19 infection and time since infection

| **Study** | **Wave Timing**  **(month-year)** | **N per wave** | ***No COVID-19** | | ***COVID-19 in last 4 weeks** | | ***COVID-19 in last 4-12 weeks** | | ***COVID-19 in last 12+ weeks** | |
| --- | --- | --- | --- | --- | --- | --- | --- | --- | --- | --- |
|  |  |  | N (%) | | N (%) | | N (%) | | N (%) | |
| MCS | May-20 | 2350 | 2030 | (86.38) | 33 | (1.4) | 188 | (8.0) | 99 | (4.21) |
| MCS | Sept-Oct 2020 | 3544 | 2535 | (71.53) | 131 | (3.7) | 39 | (1.1) | 839 | (23.67) |
| MCS | Feb-March 2021 | 4633 | 2708 | (58.45) | 54 | (1.17) | 192 | (4.14) | 1679 | (36.24) |
| ALSPAC-G1 | April 2020 | 2003 | 1709 | (85.3) | 121 | (6.0) | 124 | (6.2) | 39 | (2.5) |
| ALSPAC-G1 | May-20 | 1856 | 1538 | (82.9) | 9 | (0.5) | 139 | (7.5) | 170 | (9.2) |
| ALSPAC-G1 | December 2020 | 2646 | 2234 | (84.4) | 42 | (1.6) | 57 | (2.2) | 313 | (11.8) |
| NS | May-20 | 1678 | 1354 | (80.69) | 37 | (2.21) | 189 | (11.26) | 98 | (5.84) |
| NS | Sept-Oct 2020 | 3384 | 2400 | (70.92) | 75 | (2.22) | 19 | (0.56) | 890 | (26.3) |
| NS | Feb-March 2021 | 4069 | 2522 | (61.98) | 58 | (1.43) | 179 | (4.4) | 1310 | (32.19) |
| BCS70 | May-20 | 3436 | 2775 | (80.76) | 77 | (2.24) | 362 | (10.54) | 222 | (6.46) |
| BCS70 | Sept-Oct 2020 | 4858 | 3619 | (74.5) | 42 | (0.86) | 21 | (0.43) | 1176 | (24.21) |
| BCS70 | Feb-March 2021 | 5538 | 3739 | (67.52) | 59 | (1.07) | 178 | (3.21) | 1562 | (28.21) |
| NCDS | May-20 | 4456 | 3819 | (85.7) | 79 | (1.77) | 420 | (9.43) | 138 | (3.1) |
| NCDS | Sept-Oct 2020 | 5795 | 4671 | (80.6) | 16 | (0.28) | 12 | (0.21) | 1096 | (18.91) |
| NCDS | Feb-March 2021 | 6393 | 4791 | (74.94) | 55 | (0.86) | 142 | (2.22) | 1405 | (21.98) |
| NSHD | May-June 2020 | 1428 | 1345 | (94.79) | 0 | (0.00) | 36 | (2.12) | 47 | (3.09) |
| NSHD | Sept-Oct 2020 | 1674 | 1532 | (92.54) | 4 | (0.3) | 5 | (0.26) | 133 | (6.89) |
| NSHD | Feb-March 2021 | 1532 | 1363 | (89.1) | 3 | (0.19) | 5 | (0.21) | 161 | (10.49) |
| USoC | Apr-20 | 12,955 | 11,311 | (87.3) | 1,625 | (13) | 19 | (0.00) | 0 | (0.00) |
| USoC | May-20 | 11,730 | 10,141 | (86.5) | 167 | (1.4) | 1,422 | (12.1) | 0 | (0.00) |
| USoC | Jun-20 | 11,184 | 9,631 | (86.1) | 94 | (0.8) | 194 | (1.7) | 1,265 | (11.3) |
| USoC | Jul-20 | 10,892 | 9,278 | (85.1) | 80 | (0.7) | 258 | (2.4) | 1,276 | (11.7) |
| USoC | Sep-20 | 10,138 | 8,458 | (83.4) | 1 | (0) | 281 | (2.8) | 1,398 | (13.8) |
| USoC | Nov-20 | 9,588 | 7,794 | (81.3) | 10 | (0.1) | 258 | (2.7) | 1,526 | (15.9) |
| USoC | Jan-21 | 9,432 | 7,297 | (77.4) | 0 | (0) | 376 | (4.00) | 1,759 | (18.7) |
| USoC | Mar-21 | 10,010 | 7,567 | (75.6) | 84 | (0.8) | 72 | (0.7) | 2,287 | (22.9) |
| ALSPAC-G0 | April 2020 | 2740 | 2432 | (88.8) | 75 | (2.7) | 156 | (5.7) | 77 | (2.8) |
| ALSPAC-G0 | May-20 | 2079 | 1737 | (83.6) | <5** | (NA) | 147 | (7.1) | 191 | (9.2) |
| ALSPAC-G0 | December 2020 | 3168 | 2850 | (89) | 36 | (1.1) | 36 | (1.1) | 246 | (7.8) |
| TwinsUK | Jul-20 | 3001 | 2704 | (90.1) | 29 | (0.96) | 5 | (0.2) | 263 | (8.0) |
| TwinsUK | Oct-20 | 2799 | 2406 | (85.96) | 25 | (0.8) | 21 | (0.6) | 153 | (4.6) |
| TwinsUK | Apr-21 | 2370 | 2023 | (85.36) | 109 | (3.3) | 131 | (4.0) | 40 | (1.2) |

*After first wave of assessment values are based on “updated ever had COVID-19 infection” variable. **Collapsed due to small cell counts and potentially identifiable data in ALSPAC.

### Table S6. Descriptives for COVID-19 cases based on self-report and serology data

| Study | N with self-report and serology measures | Both self-report and serology positive | Both self-report and serology negative | Self-reported COVID-19, negative serology | No self-reported COVID-19, positive serology |
| --- | --- | --- | --- | --- | --- |
|  |  | N (%) | N (%) | N (%) | N (%) |
| MCS | 952 | 172 (18.07) | 436 (45.8) | 302 (31.72) | 42 (4.41) |
| NS | 1007 | 128 (12.71) | 554 (55.01) | 299 (29.69) | 26 (2.58) |
| BCS70 | 1927 | 162 (8.41) | 1328 (68.92) | 384 (19.93) | 53 (2.75) |
| NCDS | 2643 | 176 (6.66) | 1848 (69.92) | 573 (21.68) | 46 (1.74) |
| NSHD | 697 | 18 (2.58) | 605 (86.8) | 59 (8.46) | 15 (2.15) |
| USoC | 4,867 | 337 (8.24) | 3544 (69.97) | 851 (18.89) | 135 (2.9) |
| TwinsUK | 3137 | 263 (8.84) | 1998 (63.7) | 333 (10.6) | 543 (17.3) |

### Table S7. Covariate adjustment in GEE models

| **Study** | **Fully adjusted model covariates** | **Exceptions and notes** |
| --- | --- | --- |
| **MCS** | Sex, parent education, country of residence, ethnicity, time period of pandemic, occupational social class, prior chronic health conditions and disabilities, pre-pandemic mental health and pre-pandemic self-rated health | “Country of residence” omitted due to non-convergence in interaction model by pre-pandemic mental health. |
| **ALSPAC-G1** | Sex, education, ethnicity, time period of pandemic, parental social class, living alone status, pre-pandemic mental health and pre-pandemic life-satisfaction (wellbeing), pre-pandemic self-rated health | Pre-pandemic life-satisfaction was assessed using mental wellbeing measures |
| **NS** | Sex, education, country of residence, ethnicity, time period of pandemic, occupational social class, partnership status, prior chronic health conditions, pre-pandemic mental health and pre-pandemic life-satisfaction, pre-pandemic self-rated health | “Country of residence” omitted due to non-convergence in models with binary outcomes for satisfaction with life and interaction terms + binary outcomes |
| **BCS70** | Sex, education, country of residence, time period of pandemic, occupational social class, partnership status, prior chronic health conditions and disabilities, pre-pandemic mental health, pre-pandemic life-satisfaction, pre-pandemic self-rated health |  |
| **NCDS** | Sex, education, country of residence, time period of pandemic, occupational social class, partnership status, prior chronic health conditions, pre-pandemic mental health, pre-pandemic life-satisfaction, pre-pandemic self-rated health | “Country of residence” omitted due to non-convergence in interaction models with binary outcomes. |
| **NSHD** | Sex, education, country of residence, time period of pandemic, occupational social class, partnership status, prior chronic health conditions and disabilities, pre-pandemic mental health and pre-pandemic life-satisfaction pre-pandemic self-rated health | RQ2 - for binary life satisfaction outcome, country variable excluded due to non-convergence |
| **USoc** | Age, sex, country of residence, time period of pandemic, education, ethnicity, partnership status, pre-pandemic limiting longstanding illness, pandemic disability, pandemic self-related health, pre-pandemic psychological distress, pre-pandemic life-satisfaction. | Pre-pandemic measures were continuous/ binary depending on operationalisation of the outcome. |
| **ALSPAC-G0** | Age, sex, education, ethnicity, time period of pandemic, social class, living alone status, pre-pandemic mental health, pre-pandemic self-rated health |  |
| **ELSA** | Age (categorical), sex, time period of pandemic, education, ethnicity, partnership status, occupational social class, limiting longstanding illness, self-related health, pre-pandemic psychological distress, pre-pandemic life-satisfaction, pre-pandemic anxiety | Pre-pandemic measures were continuous/ binary depending on operationalisation of the outcome. |
| **GS** | Age (categorical), sex, time period of pandemic, education, ethnicity, partnership status, occupational social class, pre-pandemic psychological distress | Pre-pandemic measures were continuous/ binary depending on operationalisation of the outcome |
| **TwinsUK** | Age, sex, time period of the pandemic, country, education, ethnicity, partnership status, pandemic disability, pre-pandemic self-rated health, pre-pandemic psychological distress | Pre-pandemic measures were continuous/ binary depending on operationalisation of the outcome |

### Table S8. Sample descriptives

| **Characteristics (n (%))** | **MCS** | | **ALSPAC-G1** | | **NS** | | **BCS 70** | | **NCDS** | | **NSHD** | | **USoc** | | **ALSPAC-G0** | | **ELSA** | **GS** | **TwinsUK** | |
| --- | --- | --- | --- | --- | --- | --- | --- | --- | --- | --- | --- | --- | --- | --- | --- | --- | --- | --- | --- | --- |
| **Analytic sample** | 4652 | | 2498 | | 4092 | | 5545 | | 6696 | | 1721 | | 14154 | | 3258 | | 4752 | 3937 | 3137 | |
| **Sex** | | | | | | | | | | | | | | | | | | | | |
| Men | 1845 (54.97) | | 779 (31.2) | | 1530 (37.39) | | 2425 (43.73) | | 3157 (43.73) | | 810 (47.1) | | 5966 (47.7) | | 809 (24.8) | | 2127 (44.8) | 1480 (37.6) | 314 (10) | |
| Women | 2807 (45.02) | | 1719 (68.8) | | 2562 (62.61) | | 3120 (56.27) | | 3539 (56.27) | | 911 (52.9) | | 8188 (52.3) | | 2449 (75.2) | | 2625 (55.2) | 2457 (62.4) | 2823 (90) | |
| **Age group** | | | | | | | | | | | | | | | | | | | | |
| 16-29 | 5014 | | 2498 (100) | | - | | - | | - | | - | | 1573 (18.6) | | - | | 909 (19.1) | 38 (1.0) | 42 (1.3) | |
| 30-49 | - | | - | | 4092 (100) | | - | | - | | - | | 4256 (28.8) | | 57 (1.8) | | 1515 (31.9) | 781 (19.8) | 409 (13) | |
| 50-69 | - | | - | | - | | 5545 (100) | | 6696 (100) | | - | | 5951 (33.9) | | 3140 (96.4 | | 1356 (28.5) | 2295 (58.3) | 1420 (45.3) | |
| 70+ | - | | - | | - | | - | | - | | 1721 (100) | | 2374 (18.7) | | 61 (1.9) | | 972 (20.5) | 823 (20.9) | 1266 (40.4) | |
| **Education** | | | | | | | | | | | | | | | | | | | | |
| Degree | 2745 (54.75) | 1621 (64.9) | | 2088 (51.03) | | 2557 (46.11) | | 2801 (41.83) | | 444 (25.8) | | 5931 (33.3) | | 946 (29) | | 3698 (77.8) | | 1857 (47.2) | | 1552 (49.5) |
| Non-Degree | 1,927 (38.43) | 877 (35.1) | | 2004 (48.97) | | 2988 (53.89) | | 3596 (53.70) | | 1277 (74.2) | | 6543 (56.1) | | 2312 (71) | | 1054 (22.2) | | 2080 (52.8) | | 1462 (46.6) |
| *missing* | 342 (6.82) | - | | - | | - | | 299 (4.47) | | - | | 1680 (10.6) | | - | | - | | - | | 123 (3.9) |
| **Ethnicity** | | | | | | | | | | | | | | | | | | | | |
| White | 4179 (83.35) | | 2424 (97) | | 2,997 (74.28) | | - | | - | | - | | 12559 (92.4) | | 3206 (98.4) | | 4458 (93.8) | 3912 (99.4) | 3077 (98.1) | |
| Non-White | 812 (16.19) | | 74 (3) | | 1038 (25.72) | | - | | - | | - | | 1595 (7.6) | | 52 (1.6) | | 294 (6.2) | 25 (0.6) | 47 (1.5) | |
| *missing* | 23 (0.46) | | 57 | | - | | - | | - | | - | | - | | - | | - | - | 13 (0.4) | |
| **Pre-pandemic mental health** | | | | | | | | | | | | | | | | | | | | |
| Clin. symptomatic | 903 (18.01) | | 576 (23.1) | | 1057 (25.83) | | 982 (17.71) | | 855 (12.77) | | 204 (11.9) | | 2574 (20.2) | | 592 (18.2) | | 552 (11.6) | 676 (17.2) | 200 (6.4) | |
| Non symptomatic | 4111 (81.99) | | 1922 (76.9) | | 3035 (74.17) | | 4,563 (82.29) | | 5841 (87.23) | | 1517 (88.1) | | 11580 (79.8) | | 2666 (81.8) | | 4200 (88.4) | 3261 (82.8) | 2937 (93.6) | |
| **Pre-pandemic life satisfaction** | | | | | | | | | | | | | | | | | | | | |
| Satisfied | - | | - | | 3710 (90.71) | | 4821 (87.18) | | 5058 (79.07) | | - | | 10745 (72.4) | | - | | - | - | - | |
| Not Satisfied | - | | - | | 380 (9.29) | | 709 (12.92) | | 1339 (20.93) | | - | | 3409 (27.7) | | - | | - | - | - | |

### Table S9a. Mental health outcomes descriptives

|  | | | **Time 1** | | | **Time 2** | | | **Time 3** | | |
| --- | --- | --- | --- | --- | --- | --- | --- | --- | --- | --- | --- |
| **Longitudinal study** | **Mental health outcome** | **Measure** | ***N*** | **Mean score (SD)** | **High symptoms (%)** | ***N*** | **Mean score (SD)** | **High symptoms (%)** | ***N*** | **Mean score (SD)** | **High symptoms (%)** |
| **MCS** | Psychological distress | K-6 | 2,260 | 7.98 (5.09) | 18.98 | 2,813 | 8.46 (5.52) | 24.75 | 4,051 | 8.32 (5.73) | 24.66 |
|  | Depression | PHQ-2 | 2,259 | 1.84 (1.65) | 28.78 | 2,805 | 1.75 (1.70) | 27.31 | 4,050 | 1.90 (1.75) | 30.01 |
|  | Anxiety | GAD-2 | 2,257 | 1.73 (1.79) | 26.44 | 2,806 | 2.02 (1.87) | 31.70 | 4,051 | 1.97 (1.79) | 30.02 |
|  | Life satisfaction | SWL | 2,291 | 2,291 (2.21) | 35.06 | 2,834 | 6.34 (2.09) | 30.72 | 4,075 | 5.89 (2.27) | 41.24 |
| **ALSPAC- G1** | Depression | SMFQ | 1789 | 5.94 (5.36) | 16.34 | 1623 | 6.23 (5.57) | 19.70 | 2337 | 6.36 (5.97) | 20.4 |
|  | Anxiety | GAD-7 | 1788 | 6.0 (5.2) | 22.82 | 1623 | 5.76 (5.08) | 21.38 | 2343 | 6.48 (5.37) | 26.12 |
|  | Life satisfaction | WEMWBS | 1780 | 44.55 (8.28) | 29.27 | 1628 | 44.76 (8.53) | 29.91 | 2327 | 45.18 (9.31) | 29.52 |
| **NS** | Psychological distress | GHQ-12 | 1,544 | 13.46 (6.03) | 35.52 | 3,105 | 13.23 (5.74) | 30.52 | 3052 | 13.40 (5.85) | 34.40 |
|  | Depression | PHQ-2 | 1,567 | 1.41 (1.56) | 16.65 | 3,117 | 1.29 (1.63) | 18.66 | 3,569 | 1.42 (1.59) | 20.13 |
|  | Anxiety | GAD-2 | 1,570 | 1.43 (1.68) | 18.98 | 3,119 | 1.59 (1.73) | 22.48 | 3,568 | 1.51 (1.76) | 21.55 |
|  | Life satisfaction | SWL | 1,585 | 6.95 (2.06) | 20.52 | 3,131 | 6.81(2.17) | 21.96 | 3,581 | 6.53 (2.21) | 28.93 |
| **BCS70** | Psychological distress | MAL | 3,276 | 1.79 (2.00) | 18.10 | 4169 | 2.08 (2.17) | 22.73 | 4528 | 1.92 (2.11) | 21.19 |
|  | Depression | PHQ-2 | 3284 | 0.99 (1.46) | 12.65 | 4170 | 0.96 (1.50) | 12.40 | 4532 | 1.05 (1.50) | 13.26 |
|  | Anxiety | GAD-2 | 3280 | 0.99 (1.42) | 11.28 | 4170 | 1.06 (1.52) | 14.03 | 4535 | 0.97 (1.49) | 12.02 |
|  | Life satisfaction | SWL | 3303 | 7.09 (2.01) | 19.73 | 4,184 | 6.93 (2.10) | 23.38 | 4,547 | 6.84 (2.11) | 24.08 |
| **NCDS** | Psychological distress | MAL | 4,325 | 1.35 (1.82) | 12.21 | 5,507 | 1.61(1.97) | 15.87 | 5,949 | 1.50 (1.91) | 14.77 |
|  | Depression | PHQ-2 | 4,367 | 0.71 (1.32) | 8.12 | 5,502 | 0.70(1.28) | 7.90 | 5,968 | 0.87 (1.42) | 11.71 |
|  | Anxiety | GAD-2 | 4,355 | 0.77 (1.36) | 9.26 | 5,508 | 0.80(1.33) | 8.29 | 5,975 | 0.82 (1.43) | 9.50 |
|  | Life satisfaction | SWL | 4,400 | 7.36 (2.12) | 14.79 | 5,519 | 7.34(1.96) | 16.28 | 5,985 | 7.07 (2.12) | 20.80 |
| **NSHD** | Psychological distress | GHQ-12 | 1324 | 2.06 (2.51) | 20.11 | 1625 | 1.91 (2.53) | 20.88 | 1507 | 1.76 (2.84) | 18.04 |
|  | Life satisfaction | SWL | 1024 | 7.61 (1.96) | 23.19 | 1664 | 7.01 (2.19) | 34.84 | 1524 | 6.83 (2.26) | 38.85 |
| **USOC** | Psychological distress | GHQ-12 | See Table 3b below | | | | | | | | |
|  | Life satisfaction | SWL | See Table 3b below | | | | | | | | |
| **ELSA** | Depression | CESD | 4889 | 1.9 (2.1) | 19.95 | 4889 | 2.3 (2.4) | 27.08 | NA | NA | NA |
|  | Anxiety | GAD-7 | 4889 | 3.2 (4.2) | 8.52 | 4889 | 3.6 (4.5) | 10.77 | NA | NA | NA |
|  | Life satisfaction | SWL | 4889 | 7.0 (2.2) | 33.45 | 4889 | 6.9 (2.2) | 36.14 | NA | NA | NA |
| **GS** | Psychological distress | PHQ-9 and GAD-7 | 3776 | 3.56 (3.87) | 28.5 | 2784 | 3.30 (3.76) | 25.2 | 2571 | 4.17 (4.05) | 33.2 |
|  | Depression | PHQ-9 (subset) | 3836 | 2.33 (2.74) | 11.8 | 2831 | 2.33 (2.75) | 11.6 | 2612 | 2.85 (2.89) | 15.7 |
|  | Anxiety | GAD-7 (subset) | 3854 | 1.27 (1.48) | 15.9 | 2847 | 1.02 (1.36) | 11.5 | 2642 | 1.35 (1.51) | 17.6 |
|  | Life satisfaction | SWL | 3927 | 6.44 (2.27) | 44.4 | 2898 | 7.14 (2.00) | 29.6 | 2698 | 6.17 (2.22) | 47.9 |
| **ALSPAC-G0** | Depression | SMFQ | 2461 | 2.90 (3.59) | 4.06 | 2490 | 3.17 (3.89) | 5.54 | 2881 | 3.31 (3.94) | 6.0 |
|  | Anxiety | GAD-7 | 2502 | 3.41 (4.09) | 9.1 | 2529 | 3.25 (4.15) | 8.62 | 2905 | 4.22 (4.55) | 13.46 |
| **TWINS UK** | Psychological distress | HADS | 2765 | 10.2 (5.52) | 7.02 | 2623 | 10.5 (5.77) | 10.23 | 2085 | 9.04 (6.26) | 179 |
|  | Depression | HADS | 2764 | 5.28 (2.32) | 1.92 | 2620 | 5.55 (2.50) | 2.67 | 2090 | 4.46 (3.11) | 3.25 |
|  | Anxiety | HADS | 2758 | 4.91 (3.79) | 6.24 | 2617 | 4.95 (3.86) | 5.54 | 2161 | 4.66 (3.86) | 67.10 |
|  | Life satisfaction | SWL | NA | NA | NA | 2763 | 7.28 (2.01) | 25.0 | 2631 | 7.38 (1.92) | 22.3 |

NA = Not available, K-6= Kessler psychological distress scale, PHQ=Patient Health Questionnaire, GAD= Generalised anxiety disorder questionnaire, SWL= Satisfaction with Life (1 item), SMFQ= Short Mood and Feelings Questionnaire, WEMWBS = Warwick-Edinburgh Mental Wellbeing Scales, GHQ= General Health Questionnaire, MAL= Malaise Inventory, CES= Centre for Epidemiologic Studies Depression Scale, HADS= Hospital Anxiety and Depression Scale

### Table S9b. Outcome descriptive statistics – Mental health – USOC

|  | Psychological distress – GHQ12 | | | Life satisfaction - SWL 0-7 Scale | | |
| --- | --- | --- | --- | --- | --- | --- |
| Time | *N* | Mean score (SD) | High symptoms (%) | *N* | Mean score (SD) | High symptoms (%) |
| April 2020 | 12,548 | 12.62 (6.20) | 29.94 |  |  |  |
| May 2020 | 11,473 | 12.45 (6.12) | 26.70 | 11,511 | 4.80 (1.55) | 36.28 |
| June 2020 | 10,975 | 12.56 (6.23) | 25.62 |  |  |  |
| July 2020 | 10,751 | 11.79 (5.75) | 20.65 | 10,783 | 4.94 (1.58) | 31.14 |
| Sept 2020 | 10,042 | 11.89 (5.71) | 20.92 | 10,070 | 4.87 (1.51) | 33.37 |
| Nov 2020 | 9,497 | 12.84 (6.18) | 26.07 | 9,524 | 4.88 (1.52) | 32.78 |
| Jan 2021 | 9,360 | 12.82 (6.14) | 26.72 | 9,393 | 4.65 (1.53) | 40.14 |
| March 2021 | 10,038 | 12.49 (6.06) | 24.00 | 10,069 | 4.86 (1.52) | 33.05 |

### Table S10. Meta-analysed associations between COVID-19 and continuous mental health outcomes

|  | | **Mental health outcome** | | | |
| --- | --- | --- | --- | --- | --- |
|  |  | **Psychological distress** | **Depression** | **Anxiety** | **Life satisfaction** |
|  |  | Coefficient (95% CI) | Coefficient (95% CI) | Coefficient (95% CI) | Coefficient (95% CI) |
| **RQ1) COVID-19**  **(ref. no COVID-19)** | **COVID-19 (unadjusted)** | 0.13 (0.08 - 0.18) | 0.11 (0.06 – 0.17) | 0.11 (0.08 – 0.13) | -0.09 (-0.12 – -0.06) |
|  | % I2 & T^2^ | 70.6 ; 0.00 | 79.1 ; 0.00 | 26.9 ; 0.00 | 52.9 ; 0.00 |
|  | 95% PI | -0.03 – 0.29 | -0.08 – 0.30 | 0.04 – 0.16 | -0.17 – - 0.00 |
|  | **COVID-19 (adjusted)** | 0.10 (0.06 – 0.13) | 0.08 (0.05 – 0.10) | 0.08 (0.05 – 0.10) | -0.06 (-0.08 – -0.04) |
|  | % I2 & T^2^ | 42.8 ; 0.00 | 20.8 ; 0.00 | 0 ; 0.00 | 29.2 ; 0.00 |
|  | 95% PI | 0.01 - 0.17 | 0.02 - 0.12 | 0.04 - 0.10 | -0.11 – -0.00 |
| **RQ2) Duration**  **(ref. no COVID-19)** | **<4 weeks** | 0.10 (0.04 – 0.16) | 0.13 (0.05 – 0.22) | 0.07 (0.01 – 0.13) | -0.03 (-0.10 – 0.03) |
|  | % I2 | 0 | 54.3 | 0 | 0.6 |
|  | **4-12 weeks** | 0.10 (0.04 – 0.17) | 0.03 (-0.01 – 0.08) | 0.07 (0.03 – 0.11) | -0.02 (-0.09 – 0.06) |
|  | % I2 | 21.6 | 23.8 | 0 | 60.2 |
|  | **12+ weeks** | 0.10 (0.04 – 0.15) | 0.07 (0.04 – 0.10) | 0.09 (0.06 – 0.12) | -0.07 (-0.12 – -0.02) |
|  | % I2 | 62.8 | 0 | 0 | 55 |
|  | **Duration in weeks (continuous)** | -0.0001 (-0.004 – 0.002) | 0.001 (-0.002 – 0.003) | 0.001 (-0.001 – 0.002) | -0.001 (-0.003 – 0.002) |
|  | % I2 & T^2^ | 53.7 ; 0.00 | 43.5; 0.00 | 0.01 ; 0.00 | 55.36; 0.00 |
|  | 95% PI | -0.009 – 0.007 | -0.002 – 0.003 | -0.001 – 0.003 | -0.008 – 0.007 |
| **RQ3) Age stratified**  **(ref. no COVID-19)** | **16-29** | 0.08 (-0.05 - 0.20) | 0.05 (-0.00 – 0.11) | 0.04 (-0.02 – 0.10) | -0.06 (-0.11 - -0.01) |
|  | % I2 | 38.5 | 0 | 9.5 | 0 |
|  | **30-49** | 0.06 (0.01 – 0.10) | 0.04 (-0.03 – 0.10) | 0.04 (-0.04 – 0.12) | -0.03 (-0.11 – 0.05) |
|  | % I2 | 0 | 0 | 17.9 | 65 |
|  | **50-69** | 0.13 (0.10 – 0.15) | 0.10 (0.06 – 0.15) | 0.10 (0.06 – 0.13) | -0.07 (-0.11 – 0.04) |
|  | % I2 | 0 | 44.2 | 0 | 30.1 |
|  | **70+** | 0.08 (0.02 – 0.13) | 0.10 (-0.06 – 0.15) | 0.10 (-0.03 – 0.23) | -0.07 (-0.15 – 0.01) |
|  | % I2 | 0 | 71.6 | 49.8 | 0 |
| **RQ3) Interactions**  **(ref. no COVID-19)** | **Education** | 0.03 (-0.04 – 0.10) | 0.02 (-0.05 – 0.08) | 0.02 (-0.06 – 0.11) | 0.00 (-0.06 – 0.07) |
|  | % I2 | 57 | 47 | 71.4 | 46.7 |
|  | **Ethnicity** | -0.00 (-0.14 – 0.13) | -0.03 (-0.19 – 0.14) | -0.01 (-0.19 – 0.17) | 0.08 (-0.02 – 0.18) |
|  | % I2 | 0 | 24.9 | 58.7 | 0 |
|  | **Prior mental health** | 0.06 (-0.04 – 0.17) | -0.00 (-0.09 – 0.09) | 0.06 (-0.04 – 0.15) | -0.04 (-0.13 – 0.05) |
|  | % I2 | 45.2 | 49 | 19.5 | 23.8 |
|  | **Prior life satisfaction** | - | - | - | 0.04 (-0.02, 0.10) |
|  | % I2 | - | - | - | 0 |
|  | **Sex** | 0.04 (-0.01 – 0.08) | 0.03 (-0.01 – 0.07) | 0.03 (-0.02 – 0.09) | 0.00 (-0.04 – 0.05) |
|  | % I2 | 32.3 | 0 | 35.3 | 0 |
| **RQ4A) Test-confirmed vs suspected (ref. no COVID-19)** | **Suspected COVID-19** | 0.09 (0.07 – 0.11) | 0.07 (0.05 – 0.10) | 0.08 (0.05 – 0.10) | -0.05 (-0.09 - -0.02) |
|  | % I2 | 0 | 0 | 0 | 43.2 |
|  | **Test-confirmed COVID-19** | 0.11 (0.02 – 0.19) | 0.07 (-0.01 – 0.14) | 0.04 (0.00 – 0.09) | -0.00 (-0.06 – 0.05) |
|  | % I2 | 68.3 | 60.3 | 0 | 23.3 |
| **RQ4B) Serology vs self-report (ref. no COVID-19)** | **No self-report case, positive serology-** | -0.02 (-0.10 – 0.05) | -0.03 (-0.14 – 0.07) | -0.03 (-0.13 – 0.06) | 0.13 (-0.04 – 0.30) |
|  | % I2 | 0 | 0 | 0 | 0 |
|  | **Self-report case, negative serology** | 0.11 (0.06 – 0.16) | 0.13 (0.07 – 0.19) | 0.11 (0.04 – 0.17) | -0.12 (-0.23 – -0.02) |
|  | % I2 | 29.5 | 0 | 0 | 34.1 |
|  | **Self-report case, positive serology** | 0.06 (0.00 – 0.13) | 0.02 (-0.06 – 0.11) | 0.01 (-0.08 – 0.10) | -0.06 (-0.17 – 0.05) |
|  | % I2 | 10 | 0 | 9.1 | 0 |

### Table S11. Meta-analysed associations between COVID-19 and binary mental health outcomes

|  | | **Mental health outcome** | | | |
| --- | --- | --- | --- | --- | --- |
|  |  | **Psychological distress** | **Depression** | **Anxiety** | **Life satisfaction** |
|  |  | RR (95% CI) | RR (95% CI) | RR (95% CI) | RR (95% CI) |
| **RQ1) COVID-19**  **(ref. no COVID-19)** | **COVID-19 (unadjusted)** | 1.21 (1.09 – 1.36) | 1.21 (1.07 – 1.37) | 1.20 (1.09 – 1.32) | 1.11 (1.06 – 1.17) |
|  | % I2 | 93 | 90 | 84.2 | 45.7 |
|  | **COVID-19 (adjusted)** | 1.15 (1.05 – 1.25) | 1.12 (1.02 – 1.23) | 1.12 (1.04 – 1.21) | 1.09 (1.02 – 1.15) |
|  | % I2 | 88.8 | 76.8 | 63.5 | 59.9 |
| **RQ2) Duration**  **(ref. no COVID-19)** | **<4 weeks** | 1.12 (0.95 – 1.32) | 1.28 (1.02 – 1.60) | 1.02 (0.98 – 1.06) | 1.08 (0.88 – 1.33) |
|  | % I2 | 47.3 | 70.3 | 0 | 46.6 |
|  | **4-12 weeks** | 1.12 (0.98 – 1.27) | 1.03 (0.92 – 1.16) | 1.05 (0.98 – 1.12) | 1.05 (0.90 – 1.22) |
|  | % I2 | 59.3 | 42.7 | 7 | 56.1 |
|  | **12+ weeks** | 1.13 (0.98 – 1.31) | 1.05 (0.96 – 1.16) | 1.14 (1.03 – 1.26) | 1.12 (1.02 – 1.24) |
|  | % I2 | 88.4 | 58.4 | 72.4 | 59.7 |
| **RQ4A) Test-confirmed vs suspected (ref. no COVID-19)** | **Suspected COVID-19** | 1.14 (1.05 – 1.24) | 1.14 (1.04 – 1.24) | 1.13 (1.04 – 1.22) | 1.10 (1.05 – 1.15) |
|  | % I2 | 83.9 | 67.5 | 62.8 | 21.2 |
|  | **Test-confirmed COVID-19** | 1.20 (1.03 – 1.39) | 1.09 (0.93 – 1.26) | 1.04 (0.98 – 1.11) | 1.05 (0.97 – 1.13) |
|  | % I2 | 83.9 | 63.2 | 8.2 | 11 |
| **RQ4B) Serology vs self-report (ref. no COVID-19)** | **No self-report case, positive serology** | 1.01 (0.74 – 1.38) | 1.10 (0.66 – 1.83) | 1.20 (0.74 – 1.94) | 1.02 (0.79 – 1.30) |
|  | % I2 | 0 | 0 | 0 | 0 |
|  | **Self-report case, negative serology** | 1.22 (1.08 – 1.39) | 1.22 (1.01 – 1.48) | 1.28 (1.07 – 1.54) | 1.18 (1.05 – 1.33) |
|  | % I2 | 25 | 4.7 | 0 | 29 |
|  | **Self-report case, positive serology** | 1.20 (1.02 – 1.40) | 1.10 (0.84 – 1.44) | 1.12 (0.82 – 1.54) | 1.02 (0.87 – 1.18) |
|  | % I2 | 0 | 0 | 26 | 0 |

**Supplementary Figures - Forest plots**

### Figure S1. RQ1) Association between COVID-19 infection and mental health (unadjusted; continuous outcome)

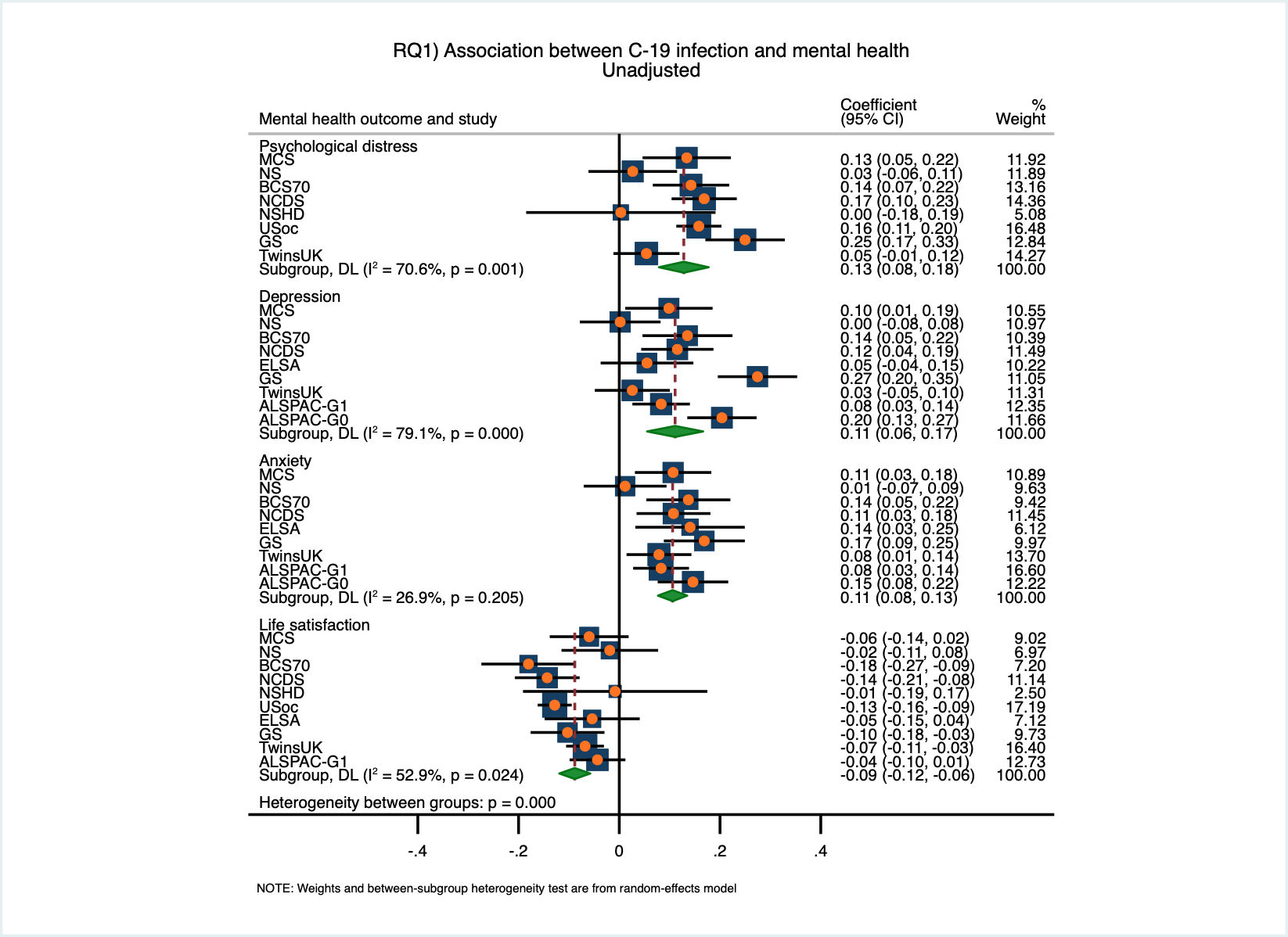

### Figure S2. RQ1) Association between COVID-19 infection and mental health (unadjusted; binary outcome)

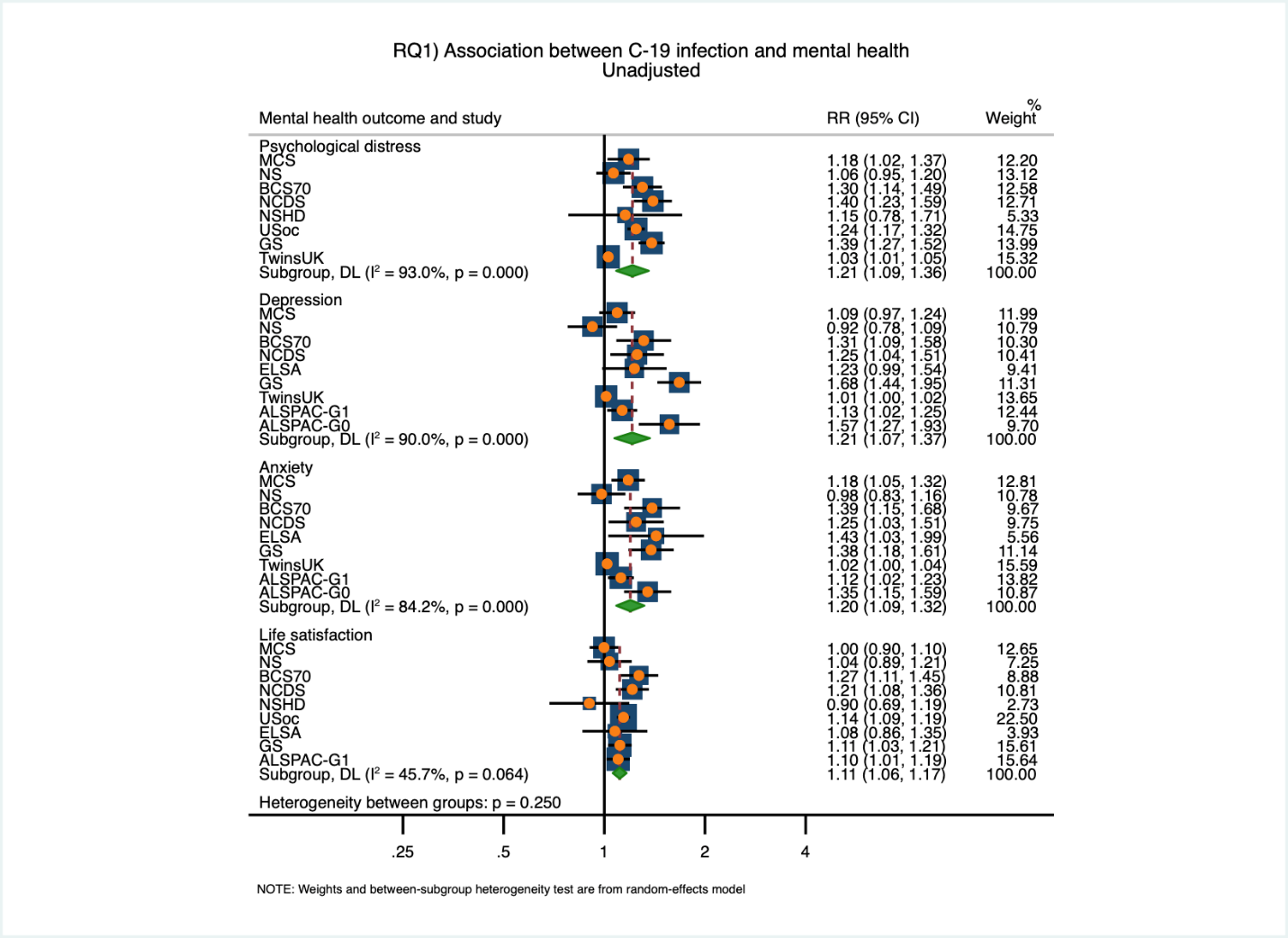

### Figure S3. RQ2) Time since COVID-19 and psychological distress (adjusted; binary outcome)

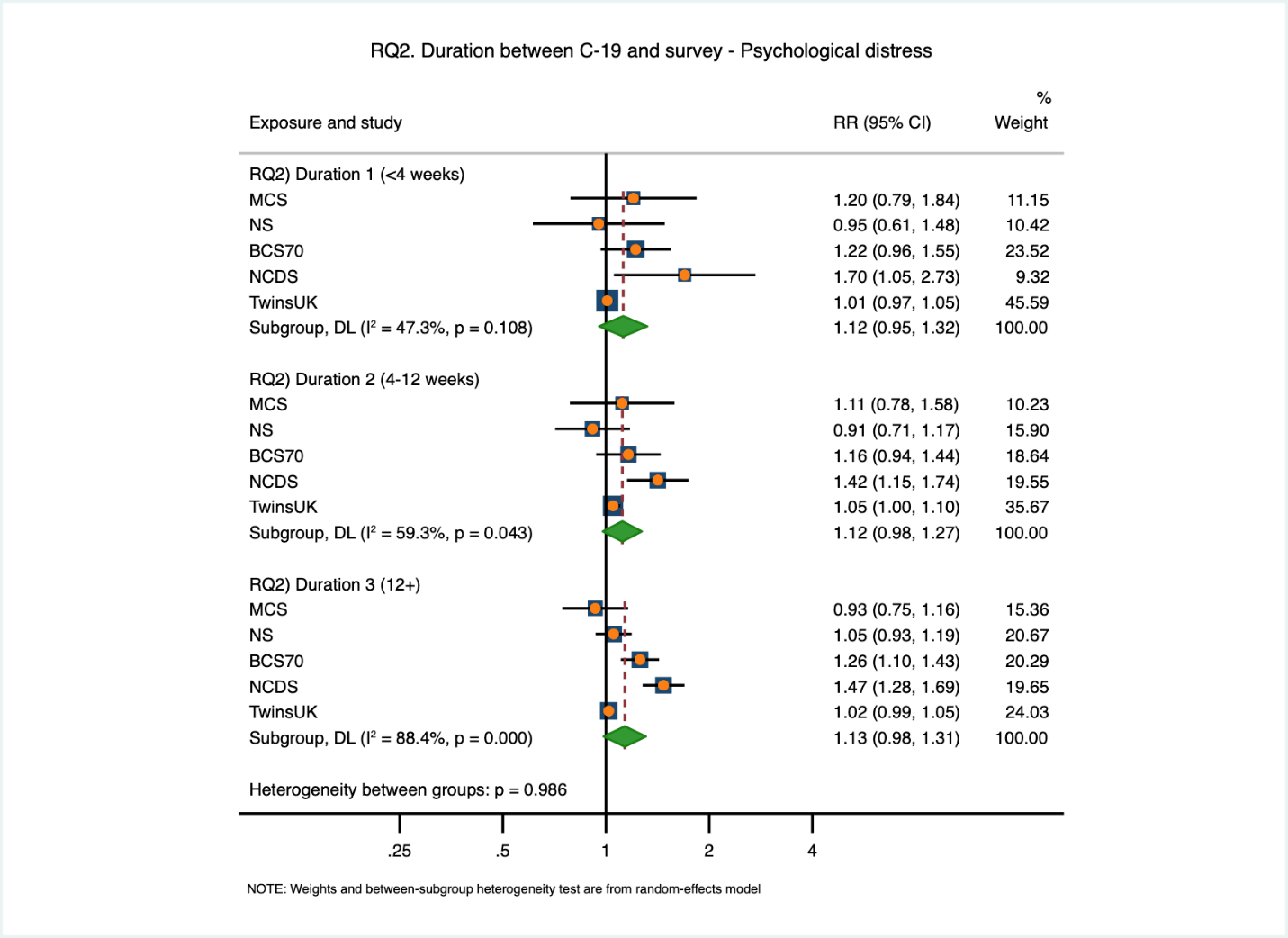

### Figure S4. RQ2) Time since COVID-19 and depression (adjusted; binary outcome)

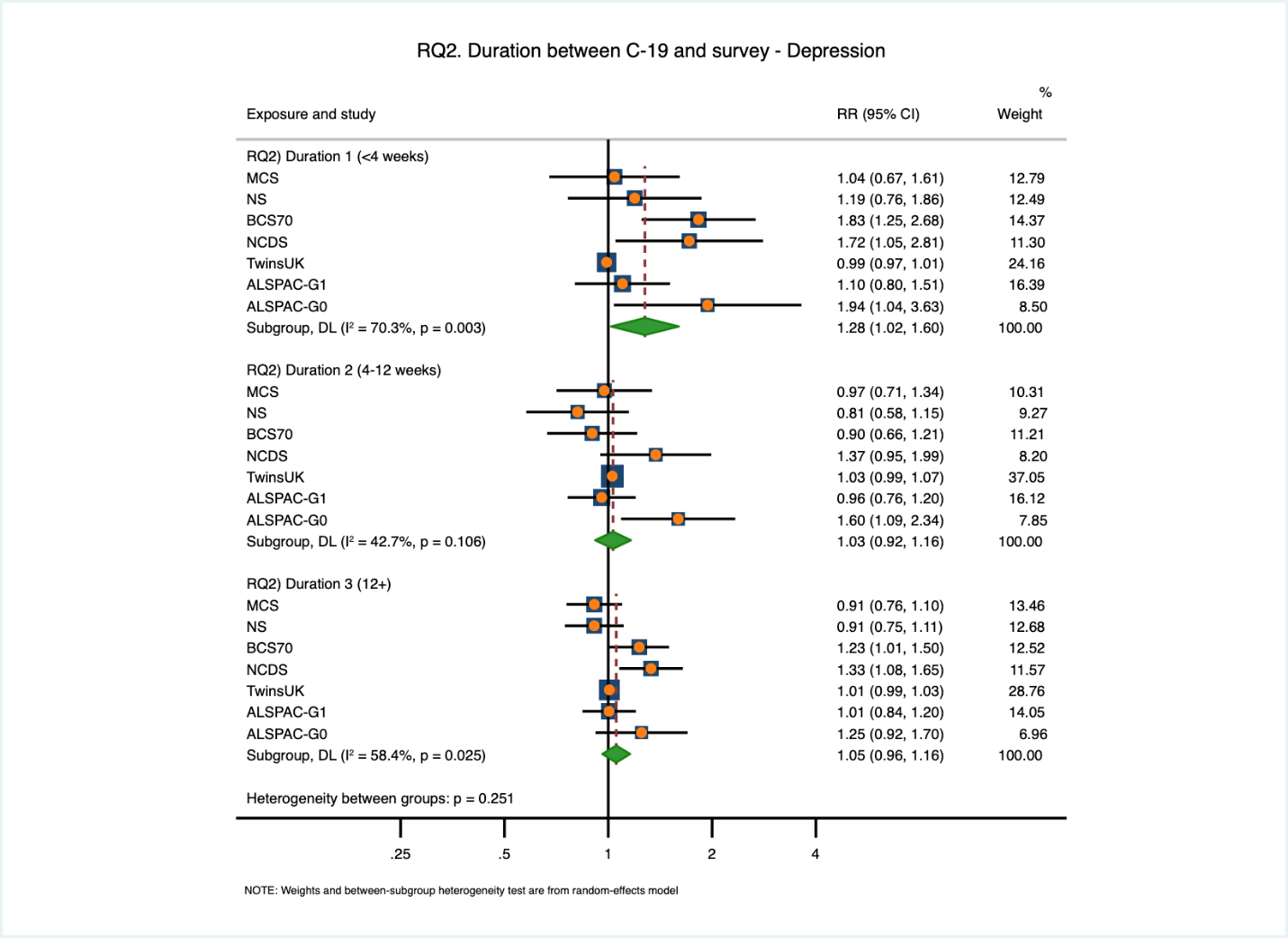

### Figure S5. RQ2) Time since COVID-19 and anxiety (adjusted; binary outcome)

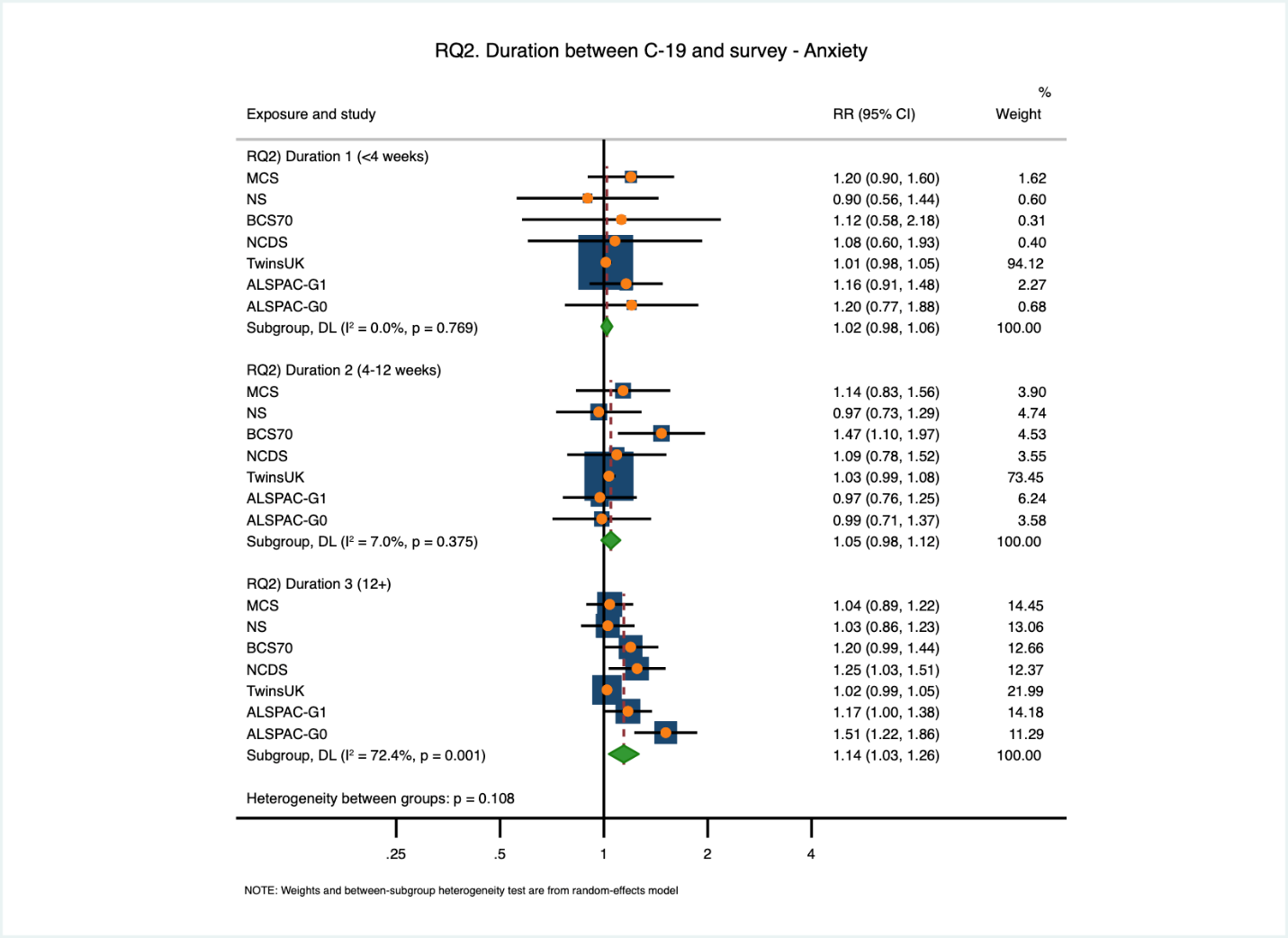

### Figure S6. RQ2) Time since COVID-19 and low life satisfaction (adjusted; binary outcome)

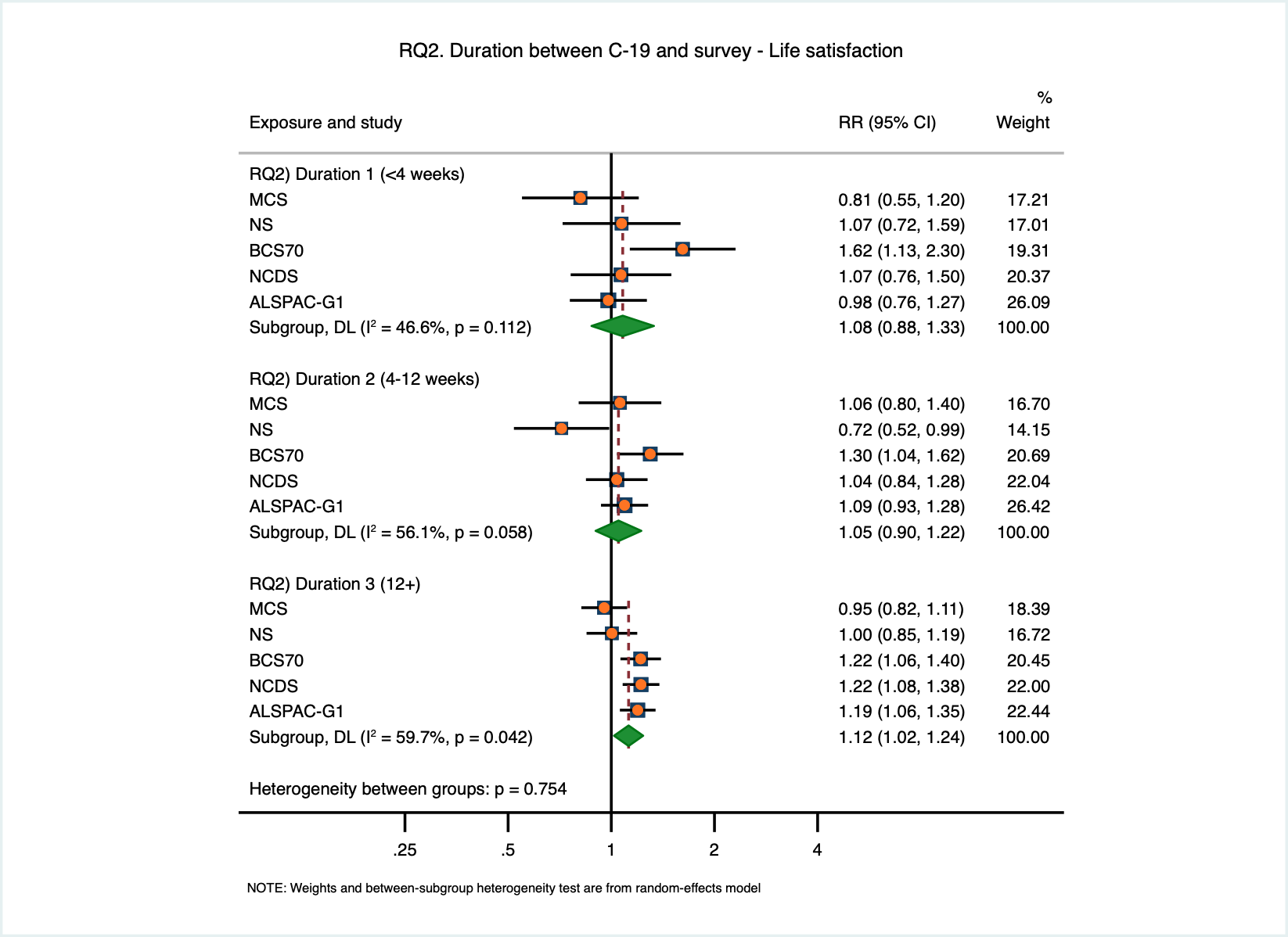

### Figure S7. RQ3) Interactions with COVID-19 - psychological distress (adjusted; continuous outcome)

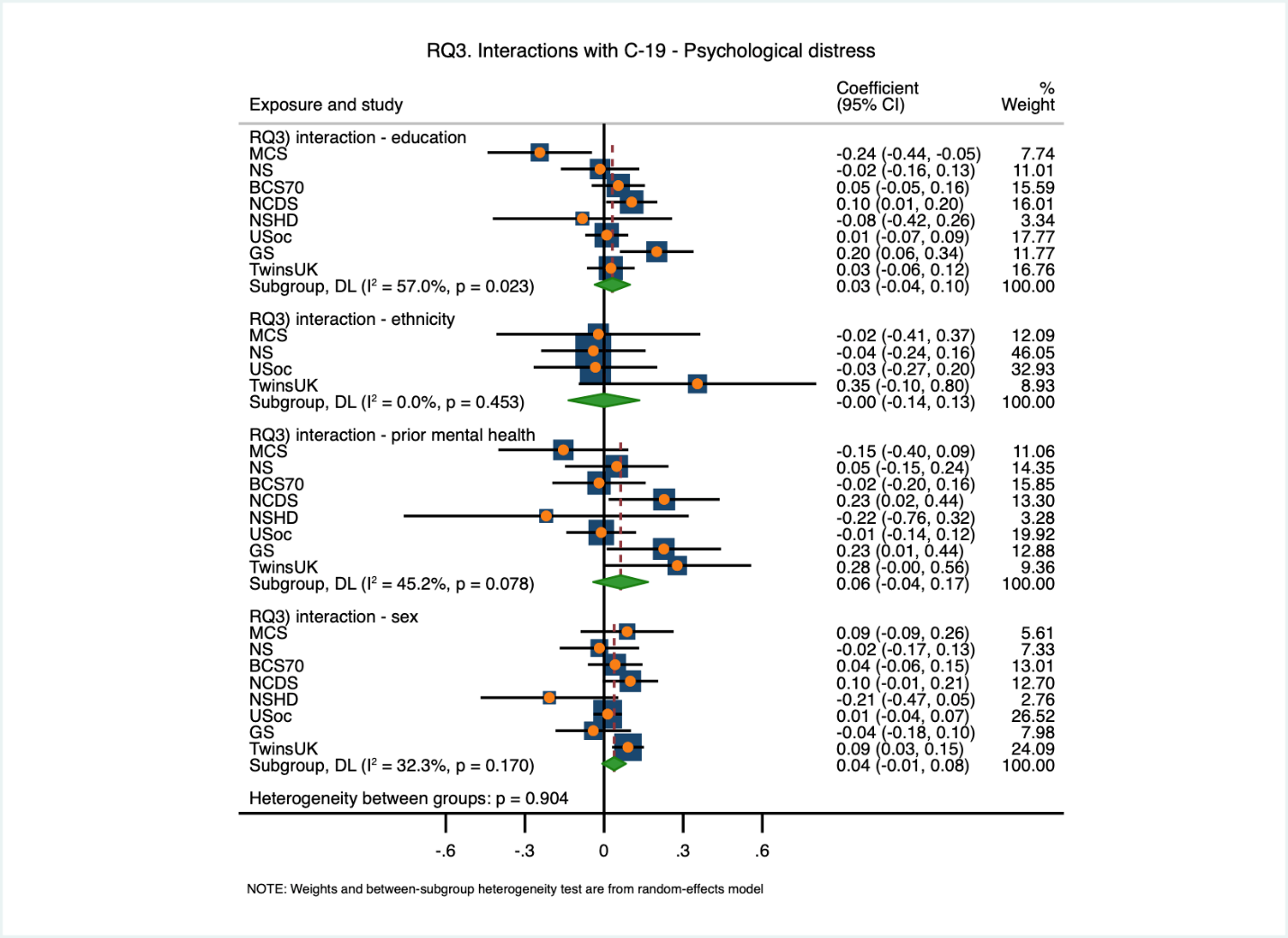

Figure S8. RQ3) Interactions with COVID-19 - depression (adjusted; continuous outcomes)

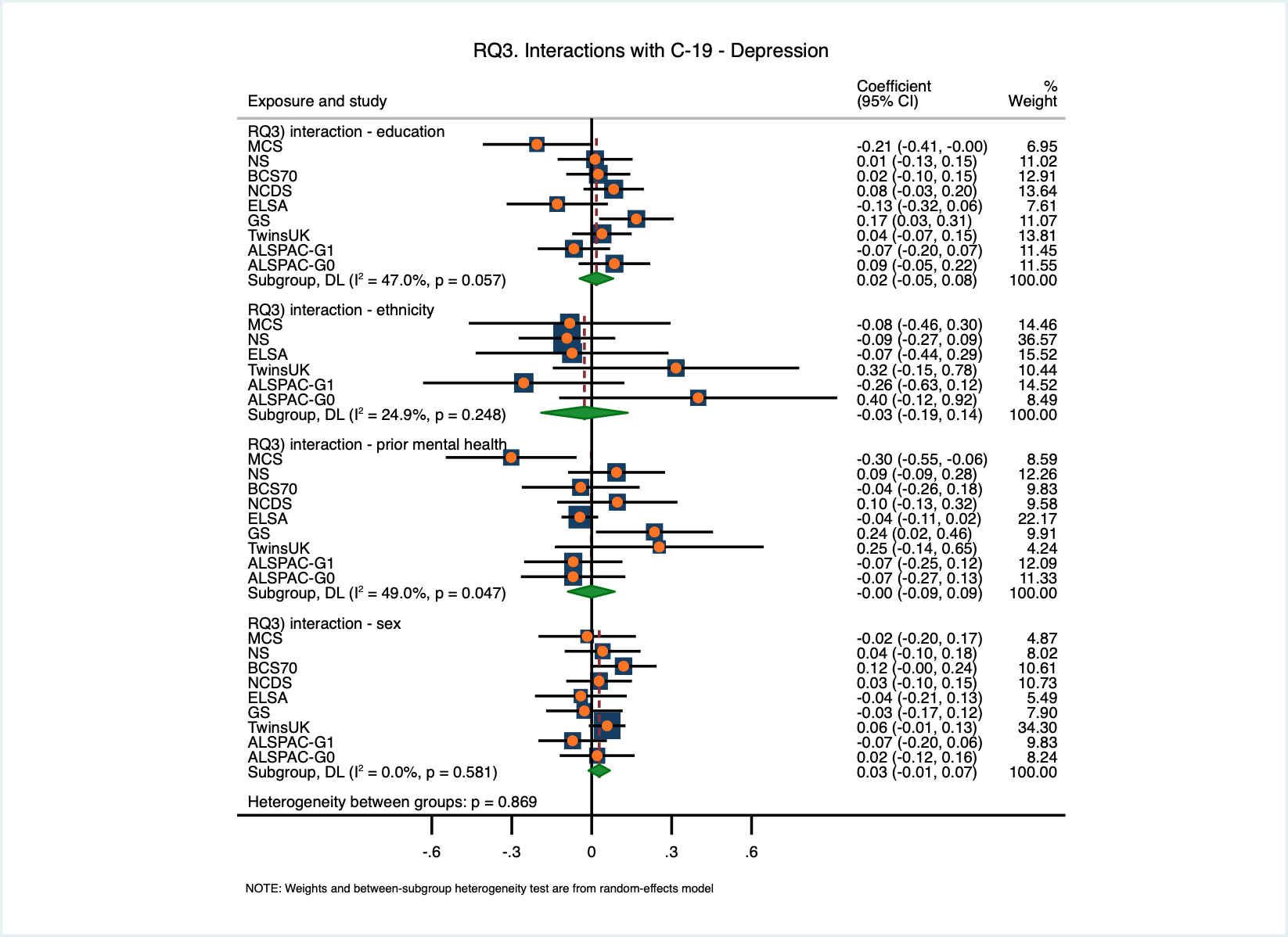

### Figure S9. RQ3) Interactions with COVID-19 - anxiety (adjusted; continuous outcomes)

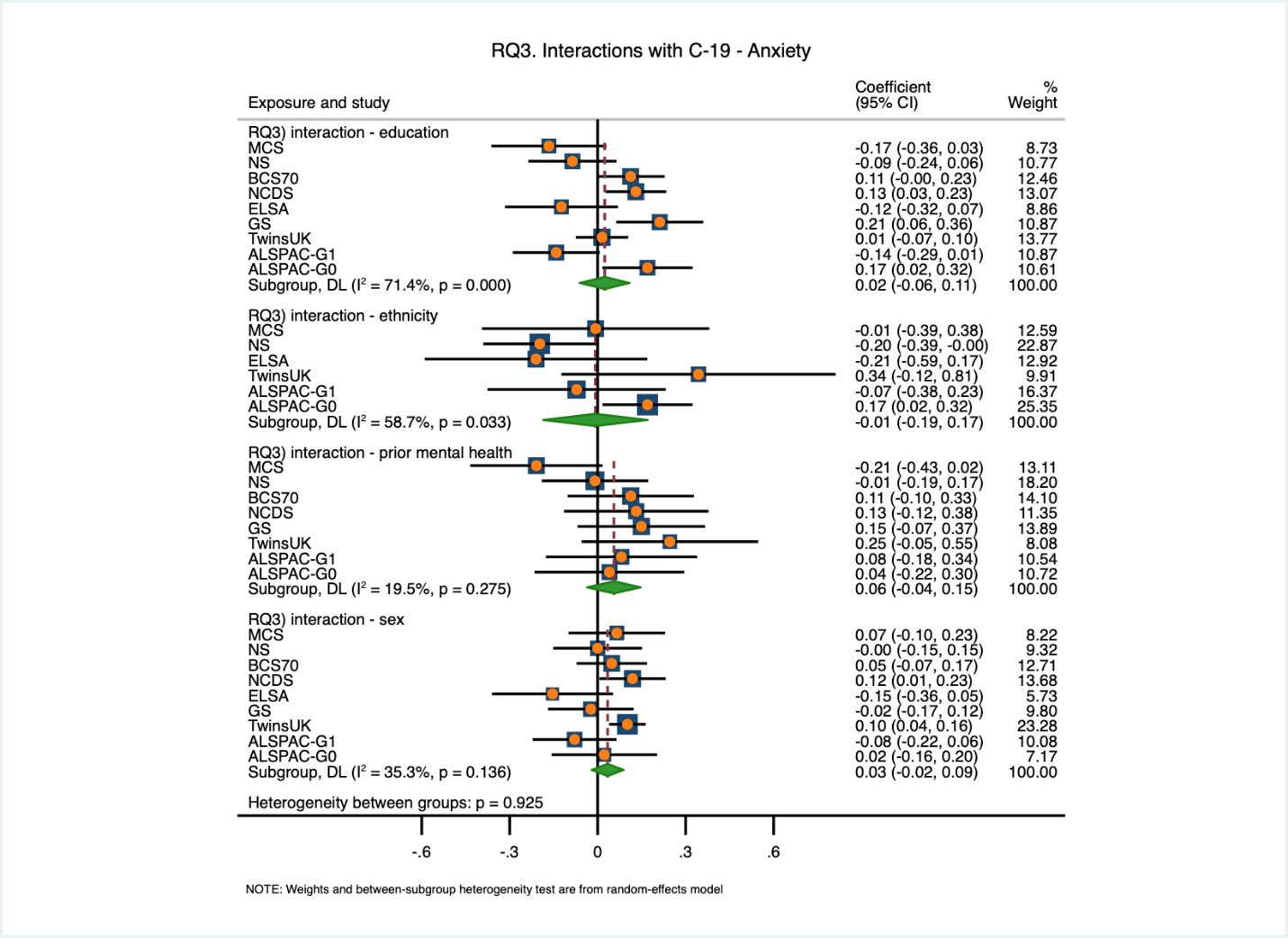

### Figure S10. RQ3) Interactions with COVID-19 - life satisfaction (adjusted; continuous outcomes)

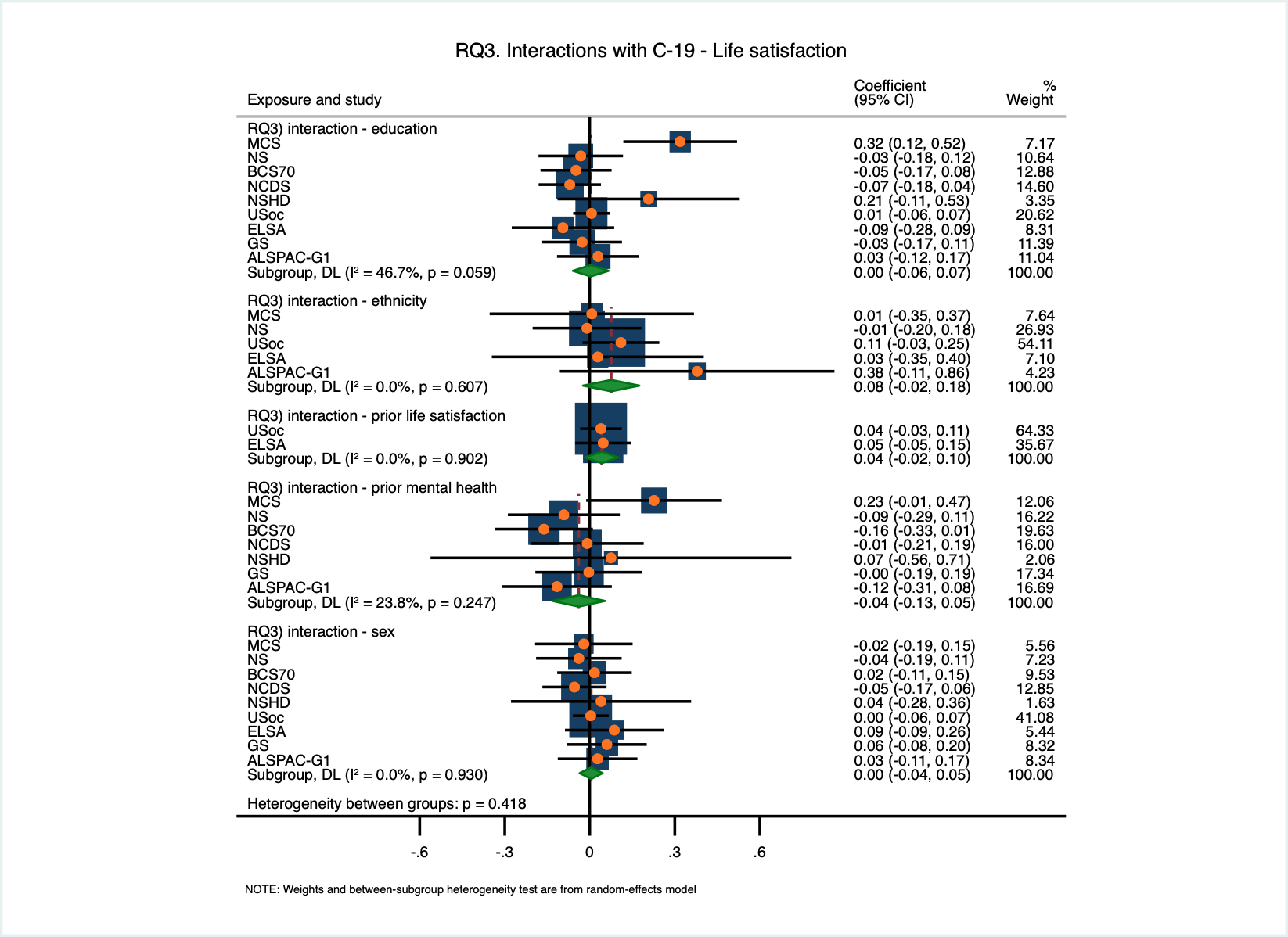

### Figure S11. RQ4A) Test-confirmed vs suspected COVID-19 and psychological distress (adjusted; binary outcome)

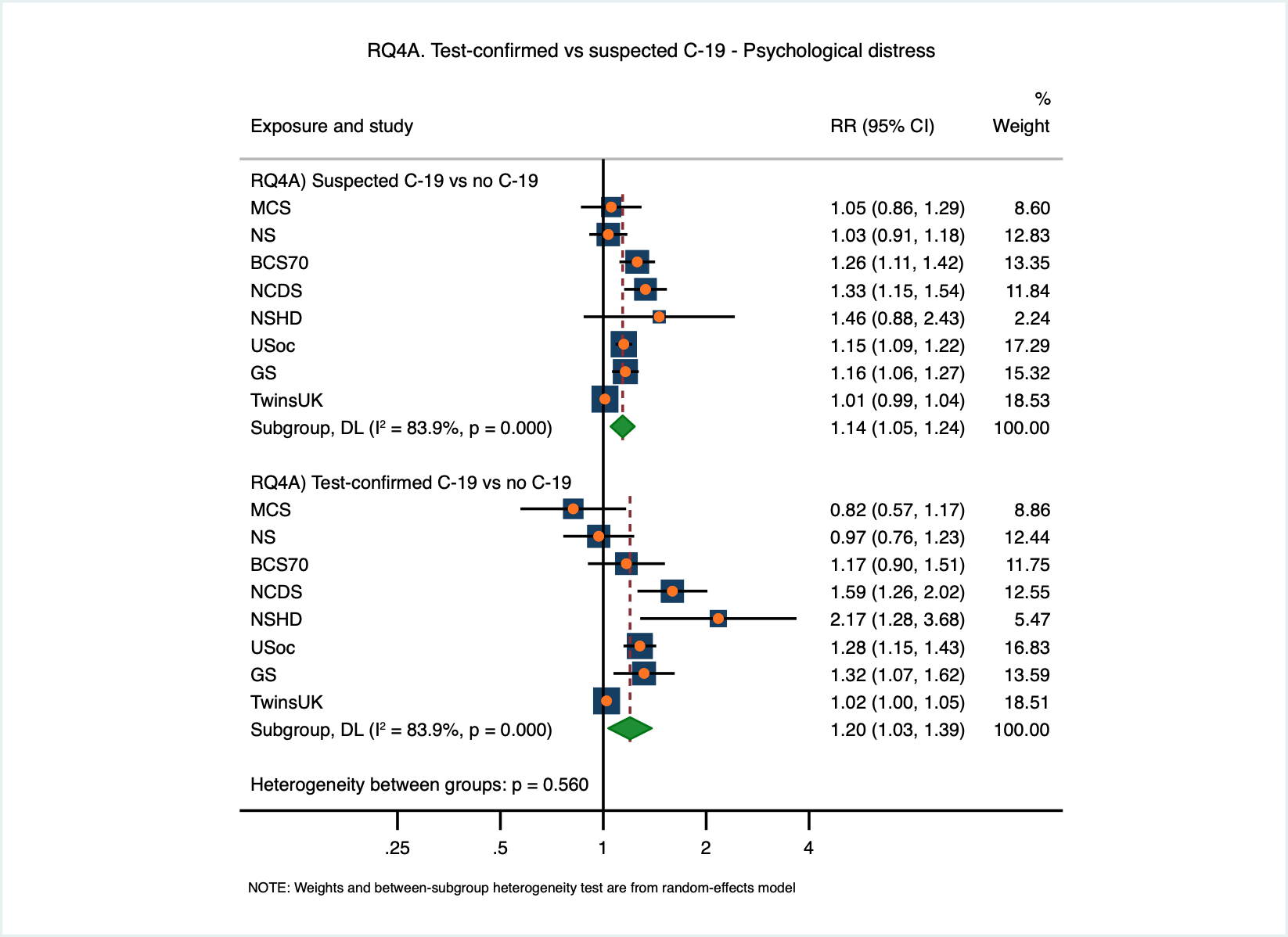

### Figure S12. RQ4A) Test-confirmed vs suspected COVID-19 and depression (adjusted; binary outcome)

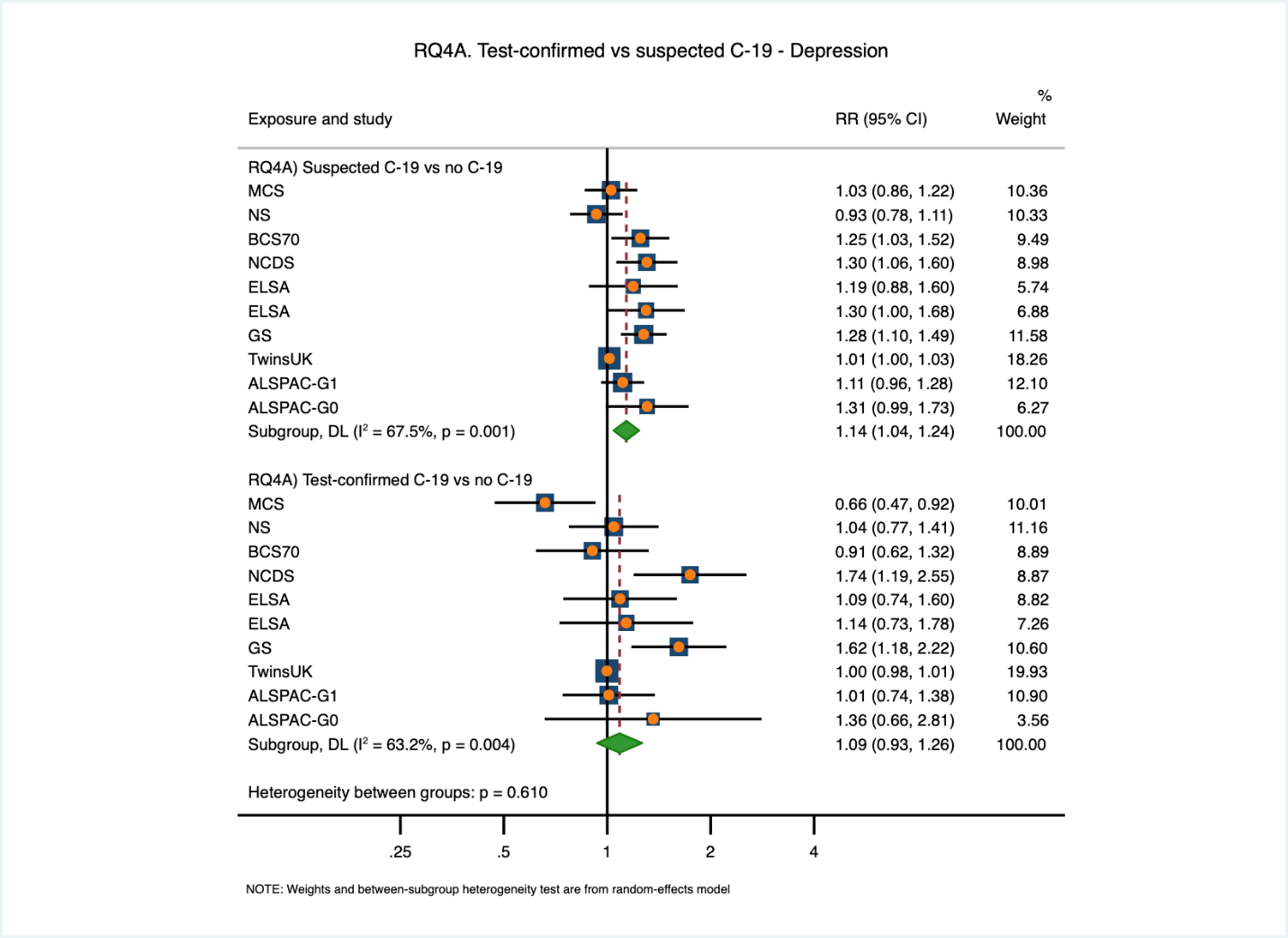

### Figure S13. RQ4A) Test-confirmed vs suspected COVID-19 and anxiety (adjusted; binary outcome)

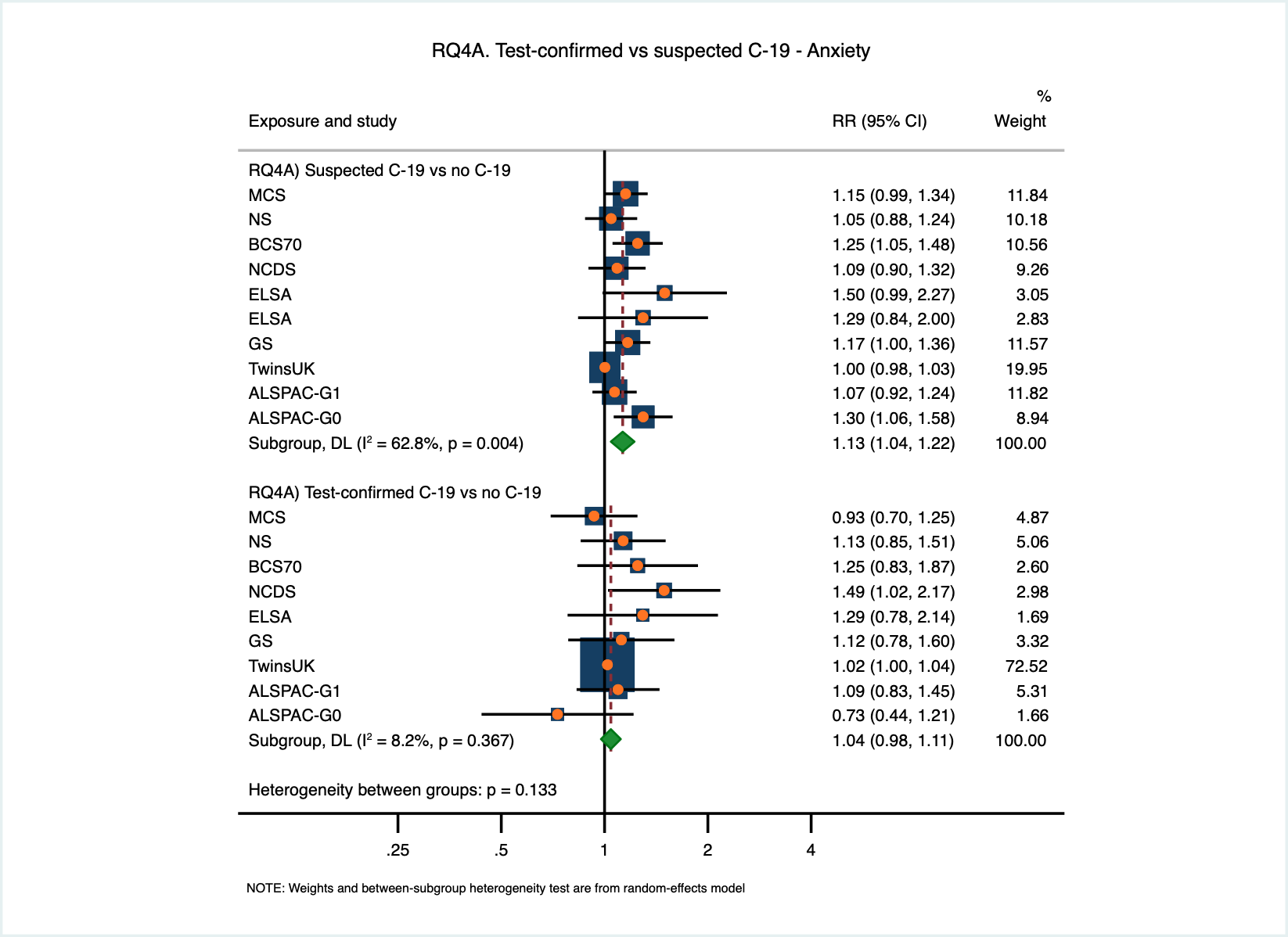

### Figure S14. RQ4A) Test-confirmed vs suspected COVID-19 and low life satisfaction (adjusted; binary outcome)

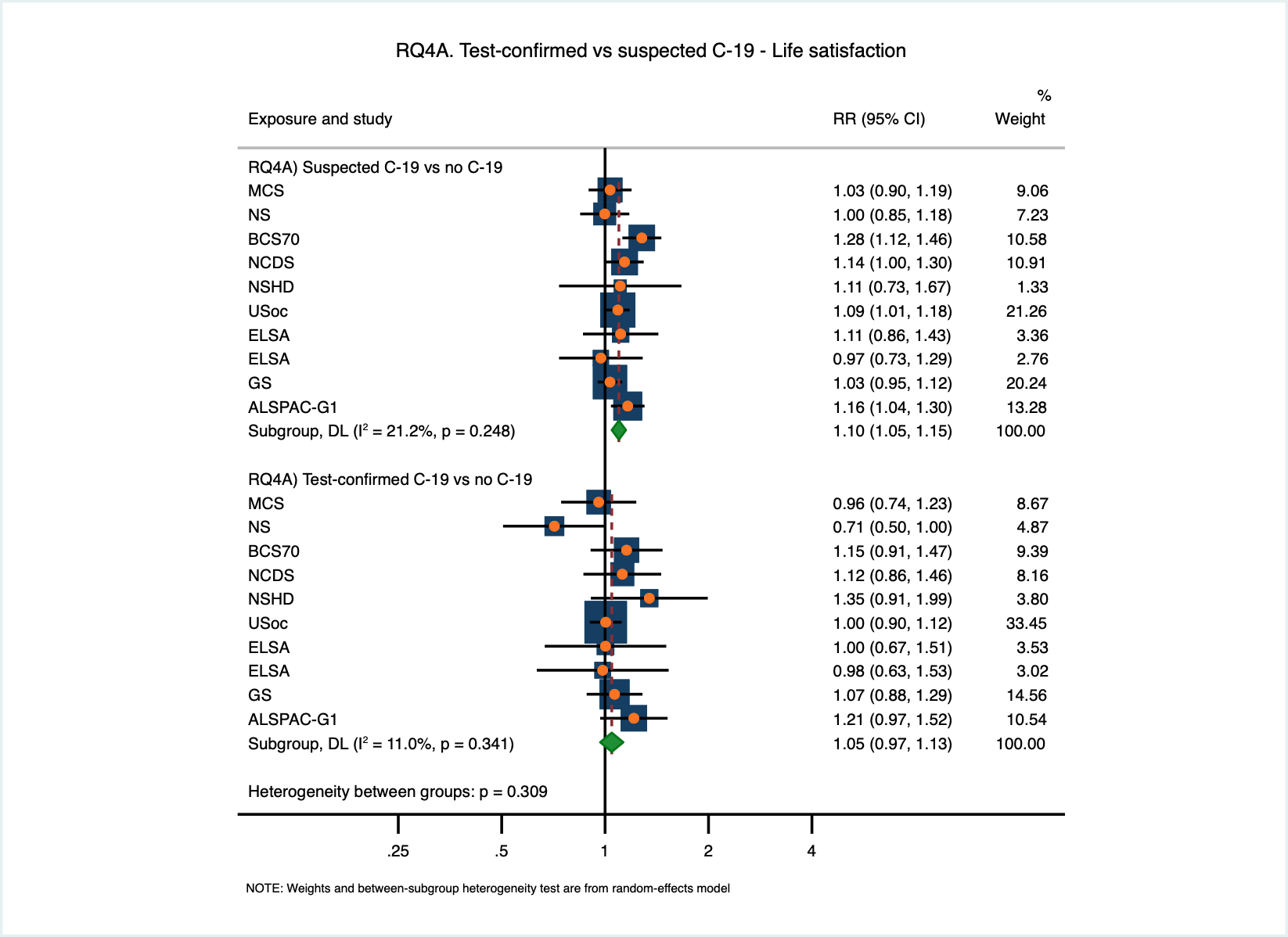

### Figure S15. RQ4B) Serology and self-report - psychological distress (adjusted; binary outcome)

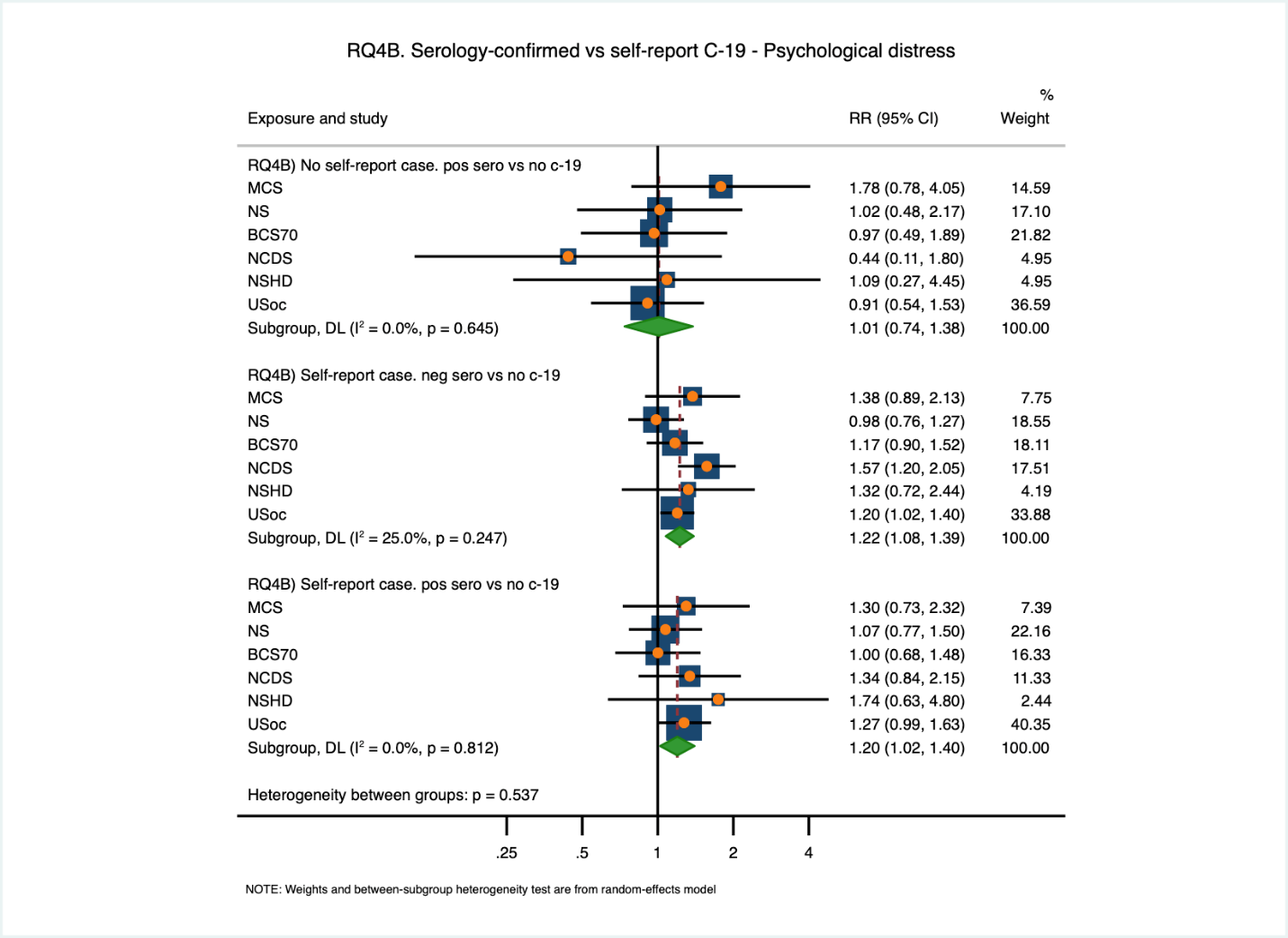

### Figure S16. RQ4B) Serology and self-report - depression (adjusted; binary outcome)

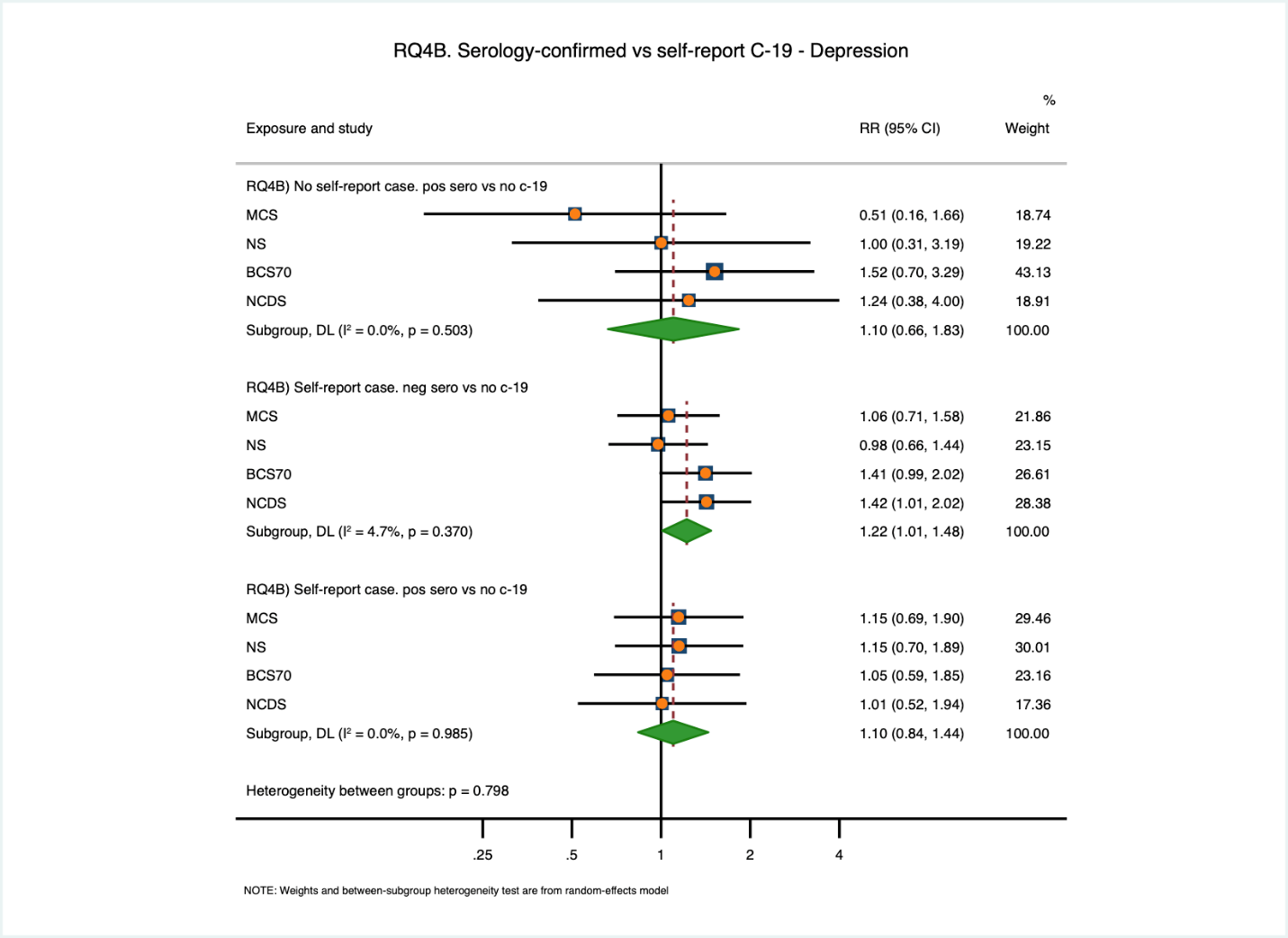

### Figure S17. RQ4B) Serology and self-report - anxiety (adjusted; binary outcome)

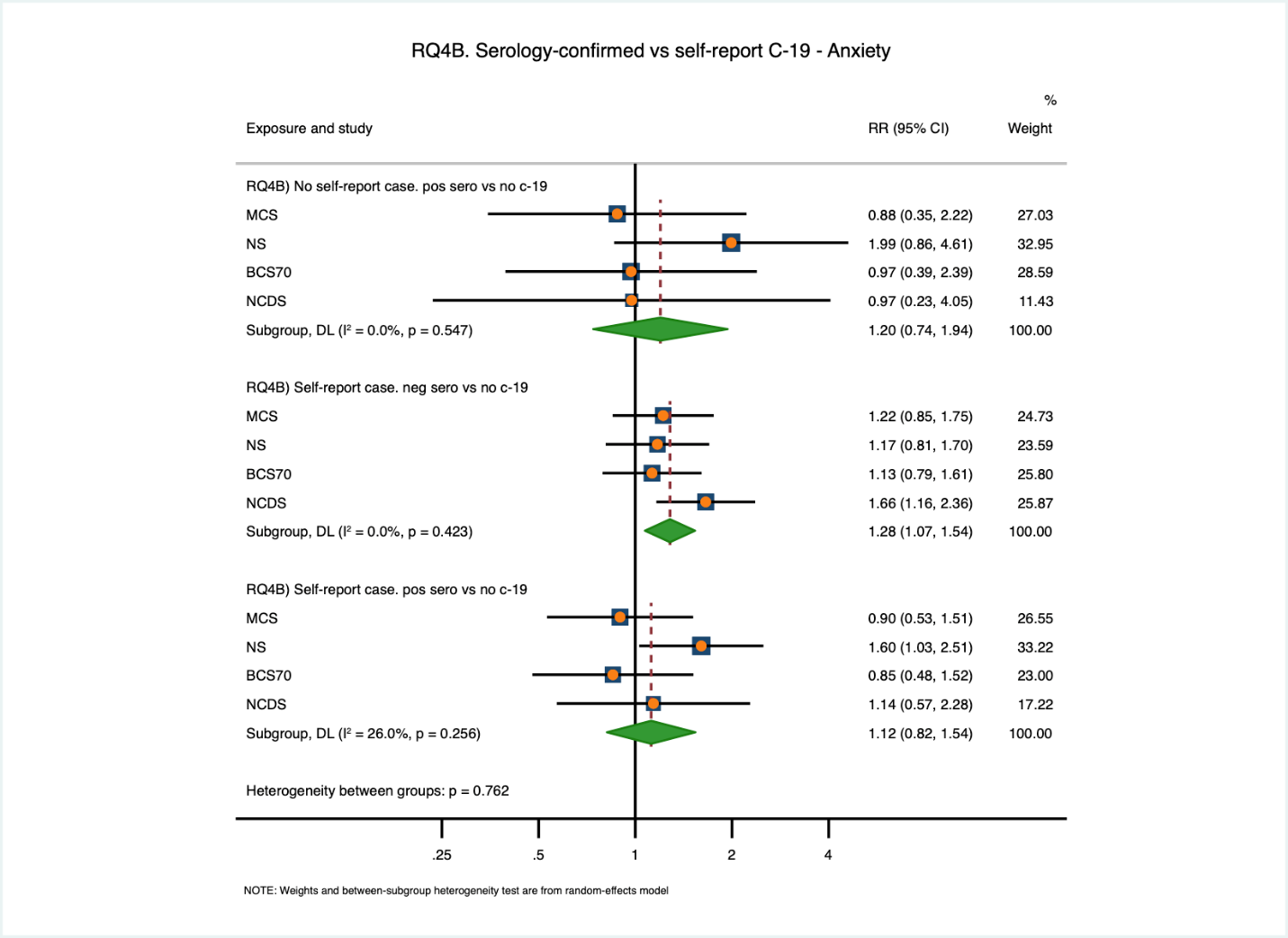

Figure S18. RQ4B) Serology and self-report - low life satisfaction (adjusted; binary outcome)

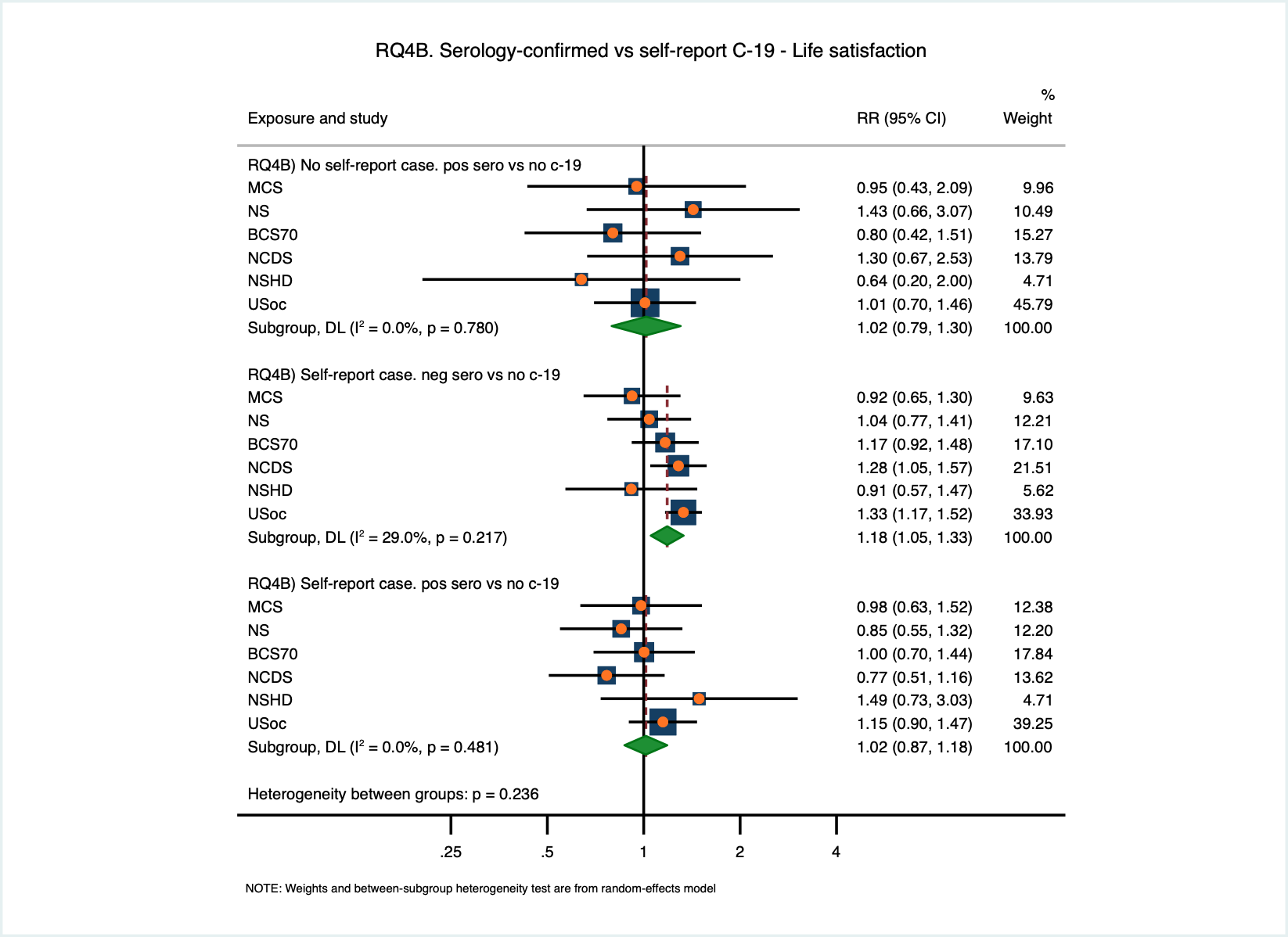

### Figure S19. Exploratory analysis – binary serology exposure – positive serology vs negative serology (continuous outcomes)

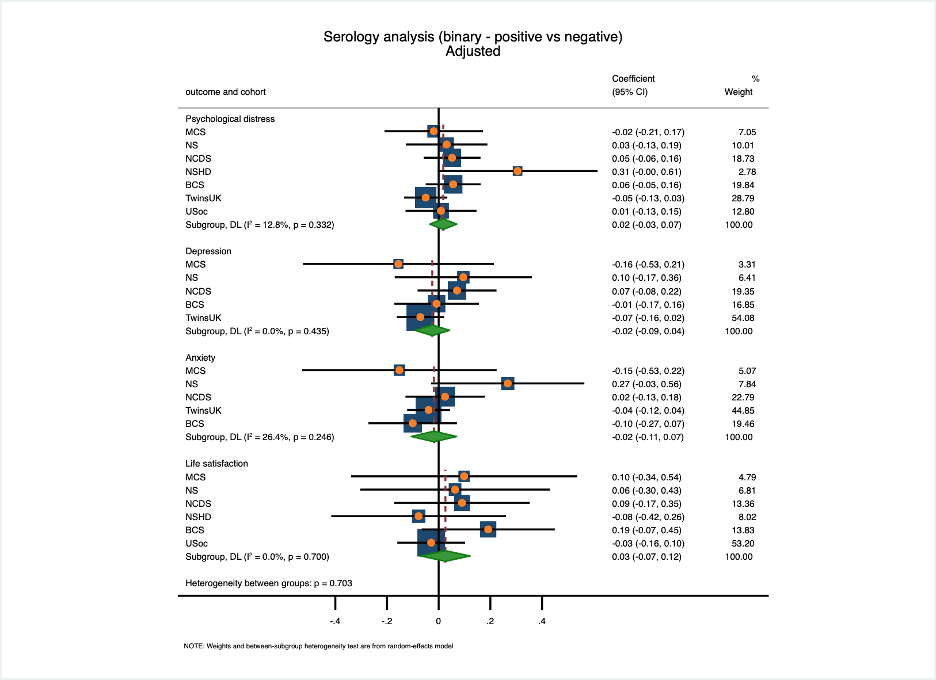

### Figure S20. RQ1 restricted to MCS, NS, BCS70, NCDS and NSHD datasets (adjusted; continuous outcome)

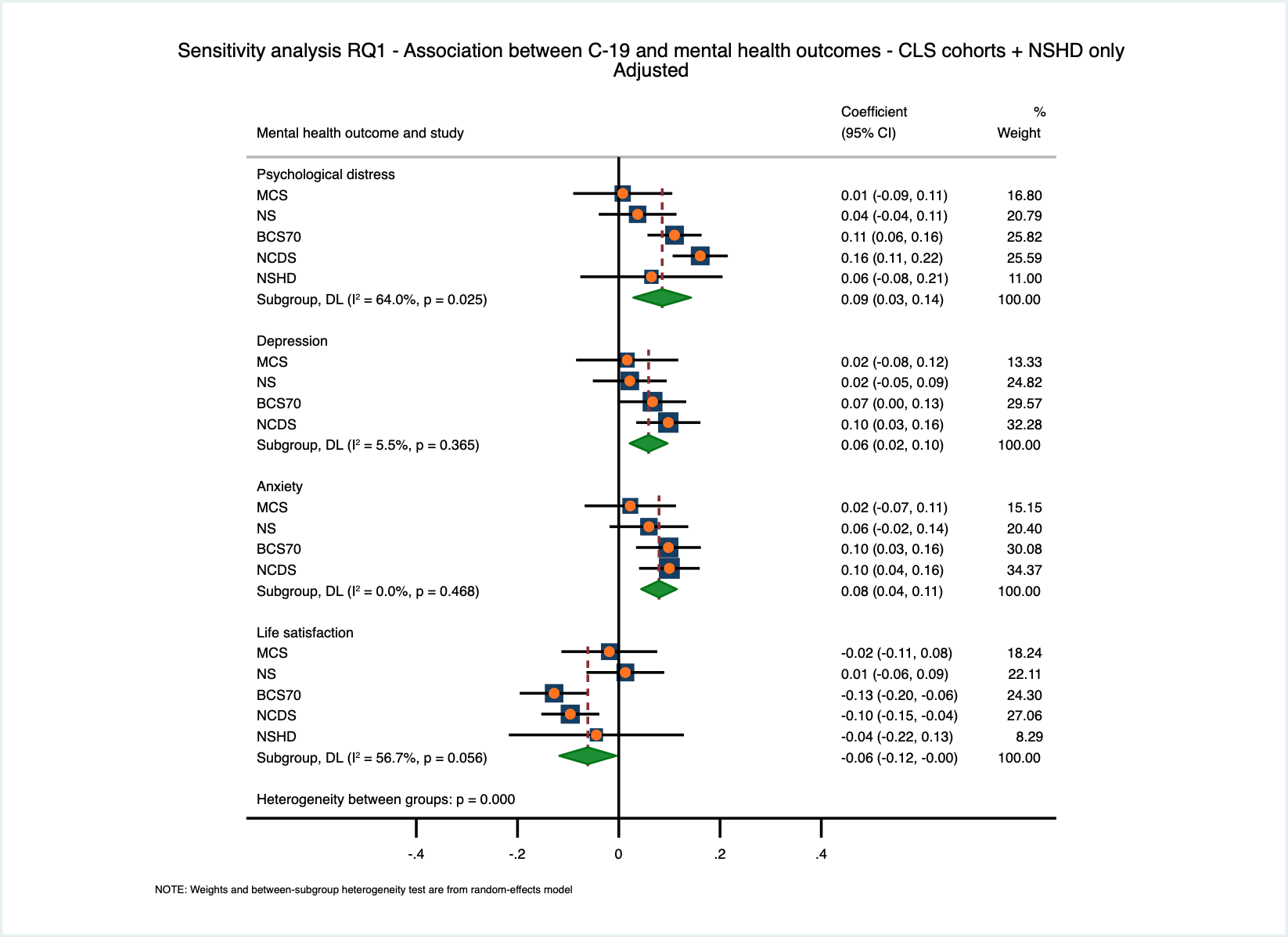

### Figure S21. Sensitivity analysis 1 - Unsure COVID-19 grouped as ‘no COVID-19’ (adjusted; continuous outcome)

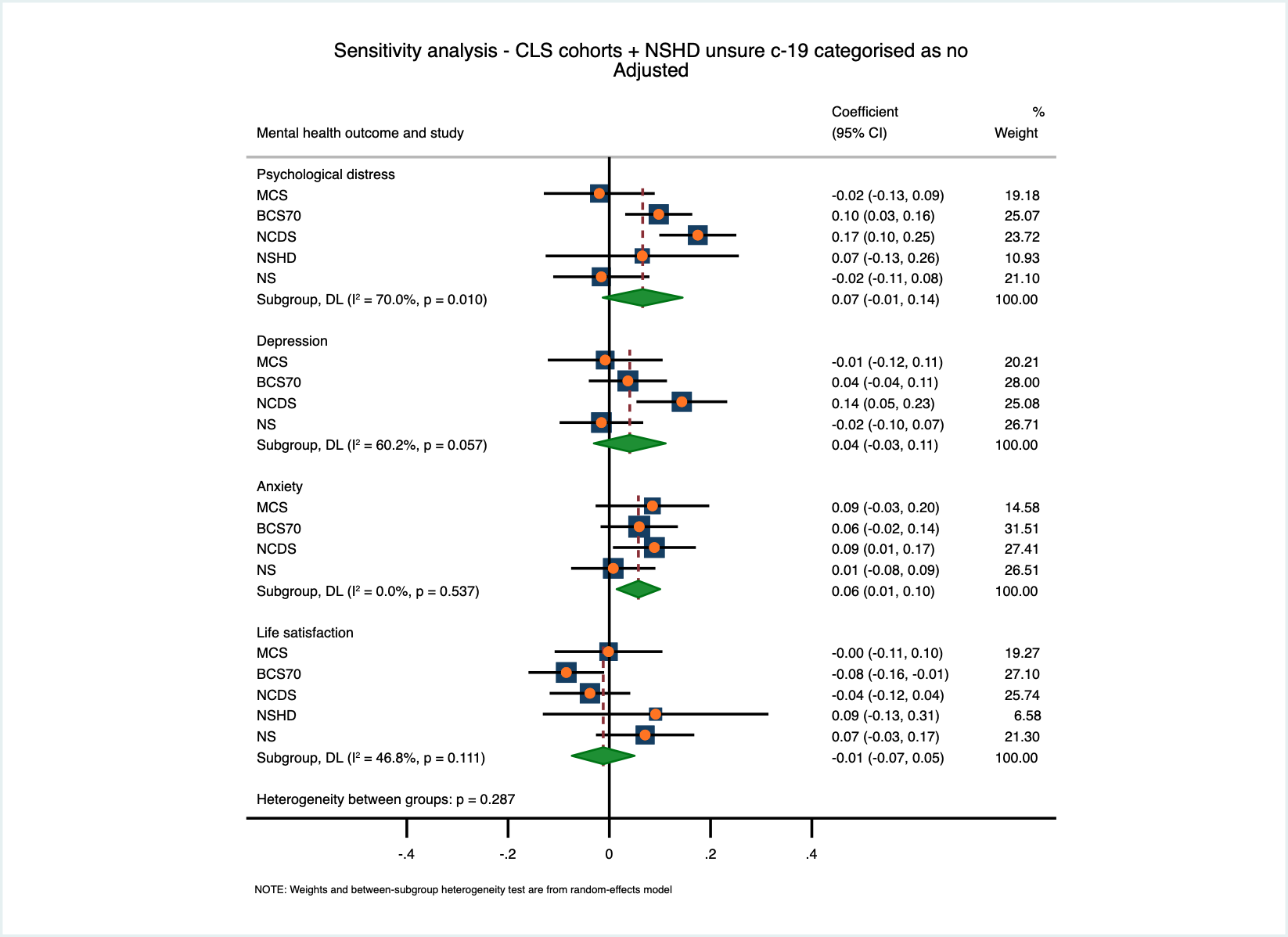

### Figure S22. Sensitivity analysis 2 - Unsure COVID-19 separate - psychological distress (adjusted; continuous outcome)

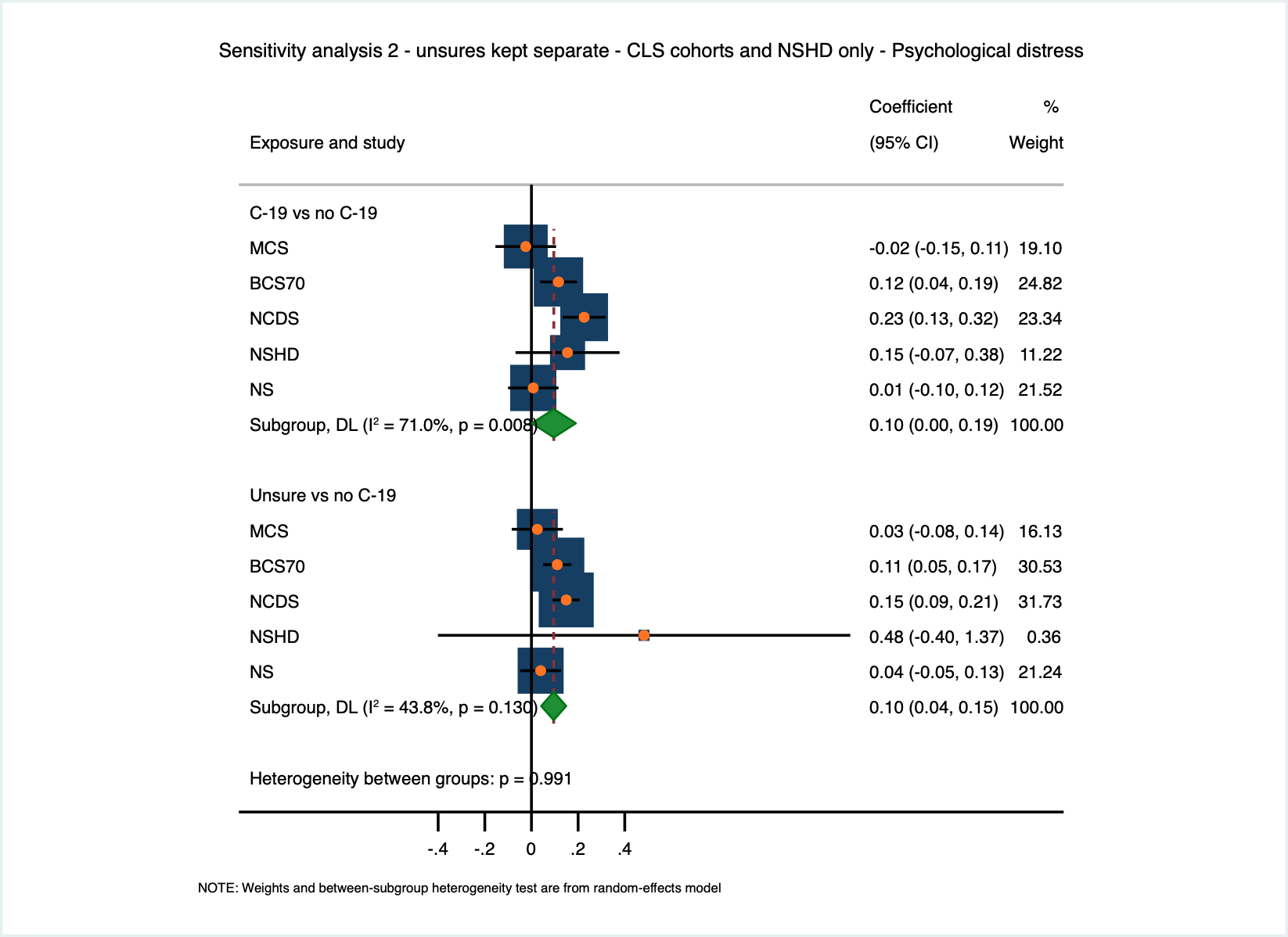

### Figure S23. Sensitivity analysis 2 - Unsure COVID-19 separate - depression (adjusted; continuous outcome)

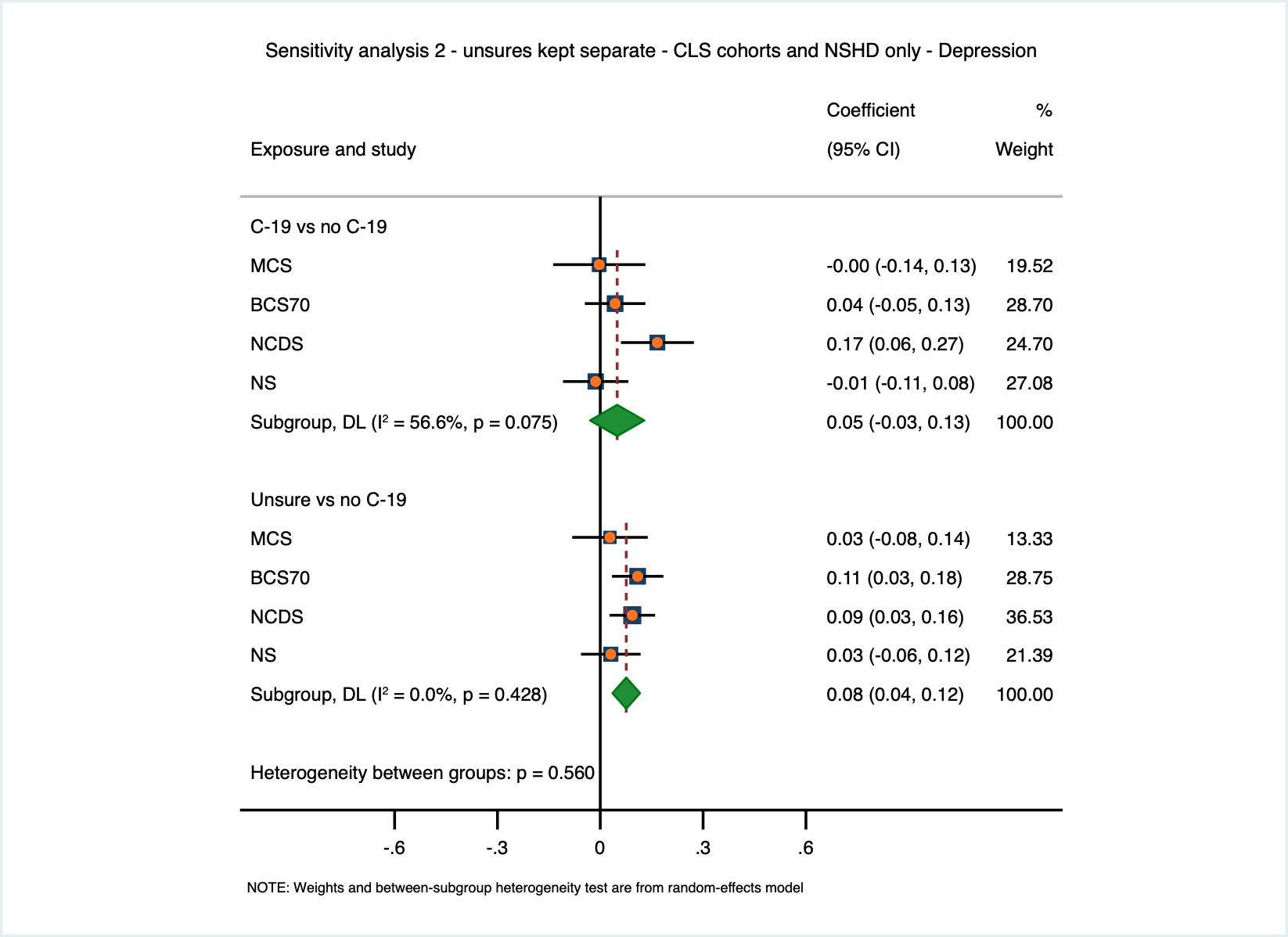

### Figure S24. Sensitivity analysis 2 - Unsure C-19 separate - anxiety (adjusted; continuous outcome)

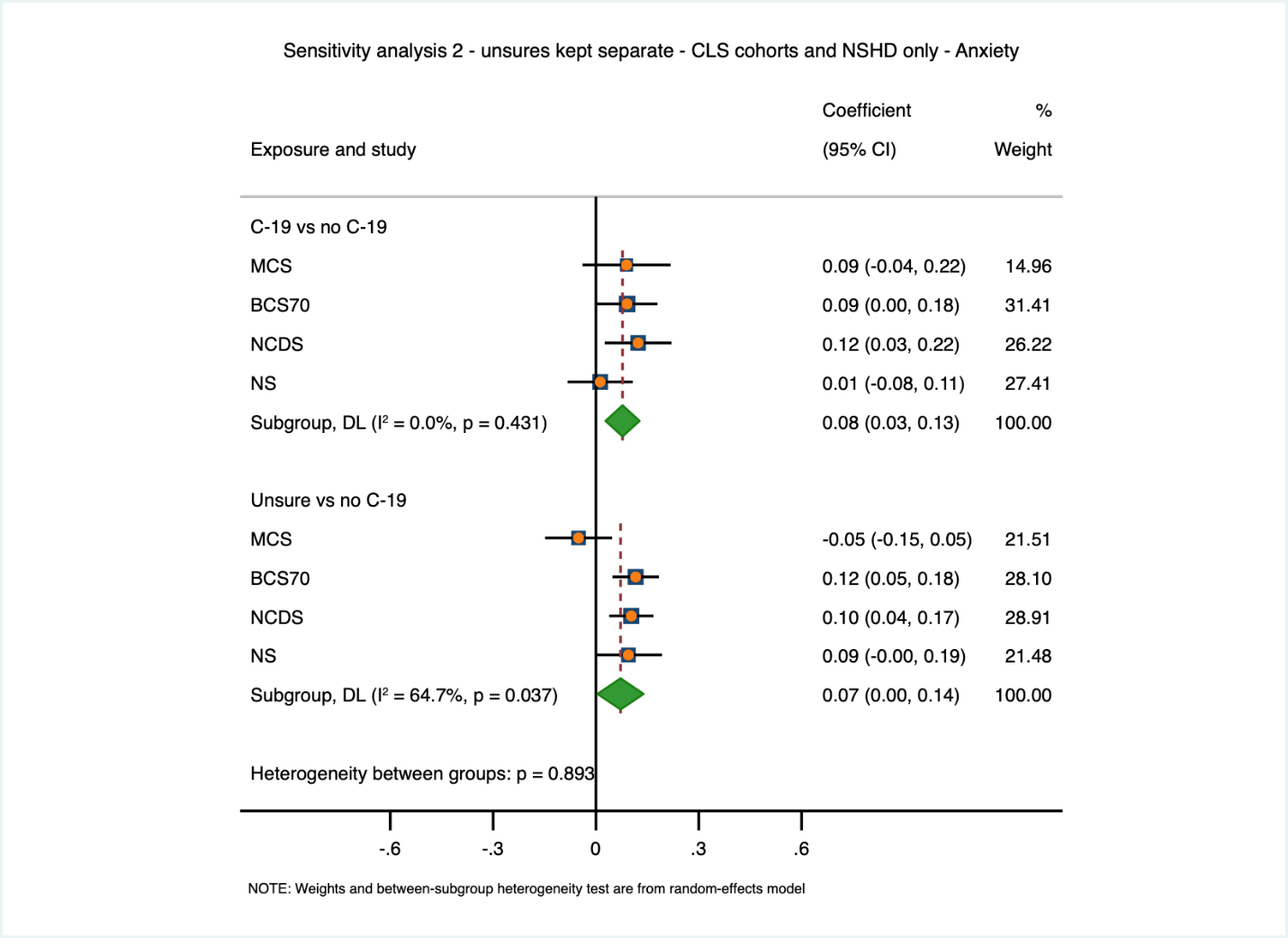

### Figure S25. Sensitivity analysis 2 - Unsure COVID-19 separate - life satisfaction (adjusted; continuous outcome)

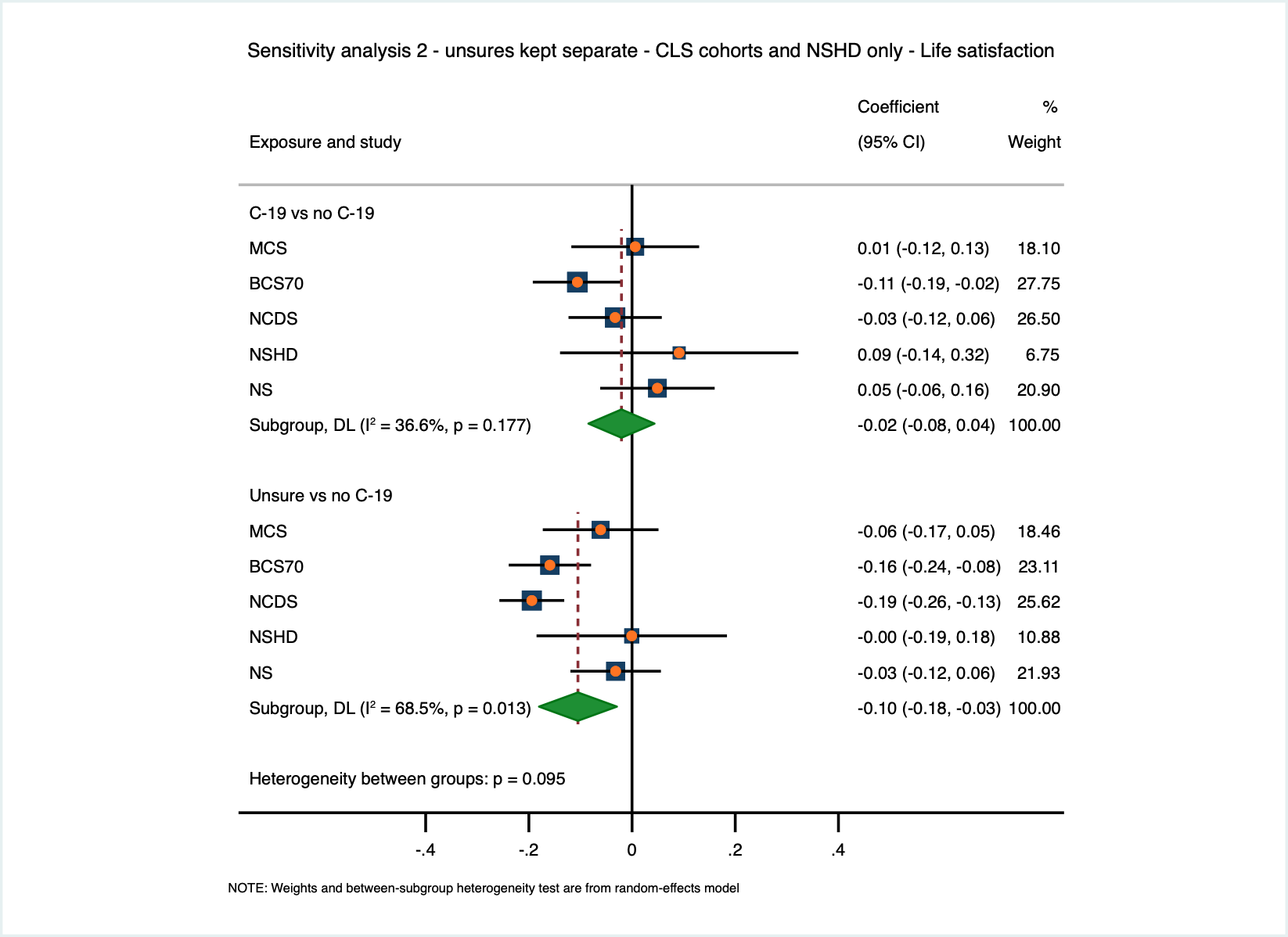

### Supplementary financial information

**ALSPAC:** The UK Medical Research Council and Wellcome (Grant Ref: 217065/Z/19/Z) and the University of Bristol provide core support for ALSPAC. A comprehensive list of grants funding is available on the ALSPAC website (<http://www.bristol.ac.uk/alspac/external/documents/grant-acknowledgements.pdf).> We are extremely grateful to all the families who took part in this study, the midwives for their help in recruiting them, and the whole ALSPAC team, which includes interviewers, computer and laboratory technicians, clerical workers, research scientists, volunteers, managers, receptionists and nurses. This work was supported by Wellcome through the Wellcome Longitudinal Population Studies COVID-19 Secretariat and Steering Group (UK LPS COVID co-ordination, Grant Ref: 221574/Z/20/Z) and supported by the Elizabeth Blackwell Institute, University of Bristol, Wellcome Trust Institutional Strategic Support Fund and Rosetrees Trust (Grant Ref: 204813/Z/16/Z; R105121). ASFK is funded by an Economics and Social Research Council (ESRC) Postdoctoral Fellowship (ES/V011650/1).

**USOC:** Understanding Society is an initiative funded by the Economic and Social Research Council and various Government Departments, with scientific leadership by the Institute for Social and Economic Research, University of Essex, and survey delivery by NatCen Social Research and Kantar Public. The Understanding Society COVID-19 study is funded by the Economic and Social Research Council (ES/K005146/1) and the Health Foundation (2076161). The research data are distributed by the UK Data Service.

**MCS, NS, BCS, NCDS, NSHD:** The Millennium Cohort Study, Next Steps, 1970 British Cohort Study and 1958 National Child Development Study are supported by the Centre for Longitudinal Studies, Resource Centre 2015-20 grant (ES/M001660/1) and a host of other co-funders. The 1946 NSHD cohort is hosted by the the MRC Unit for Lifelong Health and Ageing funded by the Medical Research Council (MC_UU_00019/1Theme 1: Cohorts and Data Collection). The COVID-19 data collections in these five cohorts were funded by the UKRI grant Understanding the economic, social and health impacts of COVID-19 using lifetime data: evidence from 5 nationally representative UK cohorts (ES/V012789/1)

**ELSA:** The English Longitudinal Study of Ageing was developed by a team of researchers based at University College London, NatCen Social Research, the Institute for Fiscal Studies, the University of Manchester and the University of East Anglia. The data were collected by NatCen Social Research. The funding is currently provided by the National Institute on Aging in the US, and a consortium of UK government departments coordinated by the National Institute for Health Research. Funding has also been received by the Economic and Social Research Council. The English Longitudinal Study of Ageing Covid-19 Substudy was supported by the UK Economic and Social Research Grant (ESRC) ES/V003941/1.

**GS:** Generation Scotland received core support from the Chief Scientist Office of the Scottish Government Health Directorates [CZD/16/6] and the Scottish Funding Council [HR03006]. Genotyping of the GS:SFHS samples was carried out by the Genetics Core   Laboratory at the Wellcome Trust Clinical Research Facility, Edinburgh, Scotland and was funded by the Medical Research Council UK and the Wellcome Trust (Wellcome Trust Strategic Award “STratifying Resilience and Depression Longitudinally” (STRADL) Reference 104036/Z/14/Z). Generation Scotland is funded by the Wellcome Trust (216767/Z/19/Z).

**TwinsUK:** TwinsUK receives funding from the Wellcome Trust (WT212904/Z/18/Z), the National Institute for Health Research (NIHR) Biomedical Research Centre based at Guy's and St Thomas' NHS Foundation Trust and King's College London. The TwinsUK COVID-19 personal experience study was funded by the King's Together Rapid COVID-19 Call award, under the projects original title ‘Keeping together through coronavirus: The physical and mental health implications of self-isolation due to the Covid-19 TwinsUK is also supported by the Chronic Disease Research Foundation and Zoe Global Ltd. The funders had no role in study design, data collection and analysis, decision to publish, or preparation of the manuscript.
